# The tryptophan catabolite or kynurenine pathway in COVID-19 and critical COVID-19: a systematic review and meta-analysis

**DOI:** 10.1101/2022.02.01.22270268

**Authors:** Abbas F. Almulla, Thitiporn Supasitthumrong, Chavit Tunvirachaisakul, Ali Abbas Abo Algon, Hussein K. Al-Hakeim, Michael Maes

**Author notes:** **Corresponding author:** Prof. Dr Michael Maes, M.D., PhD. Department of Psychiatry Faculty of Medicine, Chulalongkorn University Bangkok, 10330 Thailand. e-mail addresses.

## Abstract

Coronavirus disease 2019 (COVID-19) is accompanied by activated immune-inflammatory pathways and oxidative stress, which both may induce indoleamine-2,3- dioxygenase (IDO), a key enzyme of the tryptophan (TRP) catabolite (TRYCAT) pathway. The aim of the current study was to systematically review and meta-analyze the TRYCAT pathway status including levels of TRP and kynurenine (KYN) and IDO activity, as assessed using the KYN/TRP ratio. This systematic review was performed in December 2021 and searched data in PubMed, Google Scholar, and Web of sciences. In our meta-analysis we included 14 articles which examine TRP and TRYCATs in COVID-19 patients versus non-COVID-19 controls, and severe/critical versus mild/moderate COVID-19. Overall, the analysis was performed on 1269 individuals, namely 794 COVID-19 patients and 475 controls. The results show a significant (p <0.0001) increase in the KYN/TRP ratio (standardized mean difference, SMD=1.099, 95% confidence interval, CI: 0.714; 1.484) and KYN (SMD= 1.123, 95% CI: 0.730;1.516) and significantly lower TRP ((SMD= - 1.002, 95%CI: -1.738; -0.266) in COVID-19 versus controls. The KYN/TRP ratio (SMD= 0.945, 95%CI: 0.629; 1.262) and KYN (SMD= 0.806, 95%CI: 0.462; 1.149) were also significantly (p <0.001) higher and TRP lower (SMD= -0.909, 95% CI: -1.569; -0.249) in severe/critical versus mild/moderate COVID-19. No significant difference was detected in the kynurenic acid (KA)/KYN ratio and KA between COVID-19 patients and controls. Our results indicate increased activity of the IDO enzyme in COVID-19 and in severe/critical patients. The TRYCAT pathway is probably implicated in the pathophysiology and progression of COVID-19 and may signal a worse outcome of the disease.

**One-sentence summary:** The current meta-analysis study revealed a significant increase in peripheral blood IDO activity and kynurenine levels and a significant reduction in tryptophan in COVID-19 versus controls and in severe/critical COVID-19 versus mild/moderate COVID-19.

## Introduction

Infection with severe acute respiratory syndrome coronavirus 2 (SARS-CoV-2) may cause coronavirus disease 2019 (COVID-19) (Pakorn Sagulkoo 2022). Despite the fact that the vast majority of COVID-19 patients experience mild clinical manifestations, some patients may experience acute respiratory distress or even severe acute respiratory syndrome (SARS), which may necessitate admission to an intensive care unit (Maes, Tedesco Junior et al. 2022, Pakorn Sagulkoo 2022). Moreover, SARS has the potential to proceed to multiorgan failure and mortality, especially in the elderly and those who have concomitant illnesses such as type 2 diabetic mellitus (T2DM), hypertension, cardiovascular disease, obesity, stroke, dementia, and an increased body mass index (Mayara Tiemi Enokida Mori 2021).

COVID-19 is characterized by activated immune-inflammatory pathways and, in some cases hyperinflammation, which are accompanied by increased severity of the illness (Brosnahan, Jonkman et al. 2020, Vora, Lieberman et al. 2021). Most critically, the cytokine network is activated during SARS-CoV-2 infection, with elevated levels of many pro-inflammatory cytokines, including interleukin (IL)-1, IL-18, IL-6, tumor necrosis factor (TNF)-α, and interferon-γ (IFN-γ) (Coomes and Haghbayan 2020, Gadotti, de Castro Deus et al. 2020, Hojyo, Uchida et al. 2020, Yang, Xie et al. 2021). Mild COVID-19 may progress into SARS with pneumonia (and lowered oxygen saturation and lung lesions on chest computerized tomography scan), intravascular coagulation, multisystem failure, and death if these pro-inflammatory cytokines are overproduced during a cytokine storm (Hojyo, Uchida et al. 2020, Yang, Xie et al. 2021, Maes, Tedesco Junior et al. 2022). Profound tissue damage even extending to organ failure may be the consequence of enduring increases in IFN-γ secretion (Yin, Gribbin et al. 2005). There is now also evidence that COVID-19 is accompanied by increased production of reactive oxygen species (ROS) and ensuing oxidative damage which may contribute to severity of COVID-19 (Laforge, Elbim et al. 2020, Muhoberac 2020, Mohiuddin and Kasahara 2021). A recent protein-protein interaction network analysis based on the established gene variants associated with COVID-19 indicates the involvement of anti-viral pathways and cytokine profiles in severe COVID-19 including IFN, IL-1, IL-6 and TNF-α signaling, nucleotide-binding and oligomerization (NOD)-like receptor signaling, and Pattern Recognition Receptors – Toll-Like Receptors cascades (Pakorn Sagulkoo 2022).

During infection, increased levels of IFN-γ, IL-1β, IL-6, and ROS may induce IDO, which activates the catabolism of tryptophan (TRP) thereby lowering the levels of serum TRP and increasing levels of tryptophan catabolites (TRYCATs), including kynurenine (KYN), 3-OH-kynurenine (3HK), kynurenic acid (KA), quinolinic acid (QA), and xanthurenic acid (XA) (Maes, Leonard et al. 2011). Activation of the TRYCAT pathway protects against hyperinflammation and microbial invasion by different processes including scavenging ROS, TRP starvation, and negative immune regulatory effects (Maes, Leonard et al. 2011, Almulla and Maes 2022). Moreover, some TRYCATs, such as XA and KA have antioxidant properties (Goda, Hamane et al. 1999), while KYN, KA, XA, 3HK, and QA have negative immune regulatory effects including by attenuating IFN-γ production (Maes, Mihaylova et al. 2007, Almulla and Maes 2022). Nonetheless, following overproduction of TRYCATs, several detrimental consequences may appear, including oxidative stress, immune activation and neurotoxic effects (Dykens, Sullivan et al. 1987, Okuda, Nishiyama et al. 1998, Guidetti and Schwarcz 1999, Goldstein, Leopold et al. 2000, Santamaría, Galván-Arzate et al. 2001, Smith, Smith et al. 2009, Reyes Ocampo, Lugo Huitrón et al. 2014).

In COVID-19, some authors reported an increased activity of the TRYCAT pathway as indicated by lowered TRP and increased KYN levels and an increased KYN/TRP ratio (Shen, Yi et al. 2020, Lionetto, Ulivieri et al. 2021, Xiao, Nie et al. 2021), which reflects IDO activity (Maes and Anderson 2021). **Figure 1** shows the possible role of the TRYCAT pathway in COVID-19. Probably, the IDO enzyme, which is the first and rate-limiting enzyme of the TRYCAT pathway, is induced in COVID-19 by increased levels of IFN-γ, IL-1, IL-6, TNF-α, and ROS (Maes, Leonard et al. 2011). Vyavahare et al. systematically reviewed the alterations of KYN and TRP in COVID-19 and suggested probable implications of the TRYCAT pathway in bone and muscle tissue damage (Vyavahare, Kumar et al. 2021). Moreover, it was proposed that stimulation of the aryl hydrocarbon receptor (AhR) by coronaviruses and IDO-induced KYN levels may cause the “systemic aryl hydrocarbon receptor activation syndrome” (SAAS), which aggravates hyperinflammation, hypercoagulation, and organ injuries (Turski, Wnorowski et al. 2020). Overall, TRYCAT pathway activation in COVID-19 may worsen the illness and probably decrease the patient’s recovery potential (Robertson, Gostner et al. 2020, Lionetto, Ulivieri et al. 2021).

**Figure 1:**
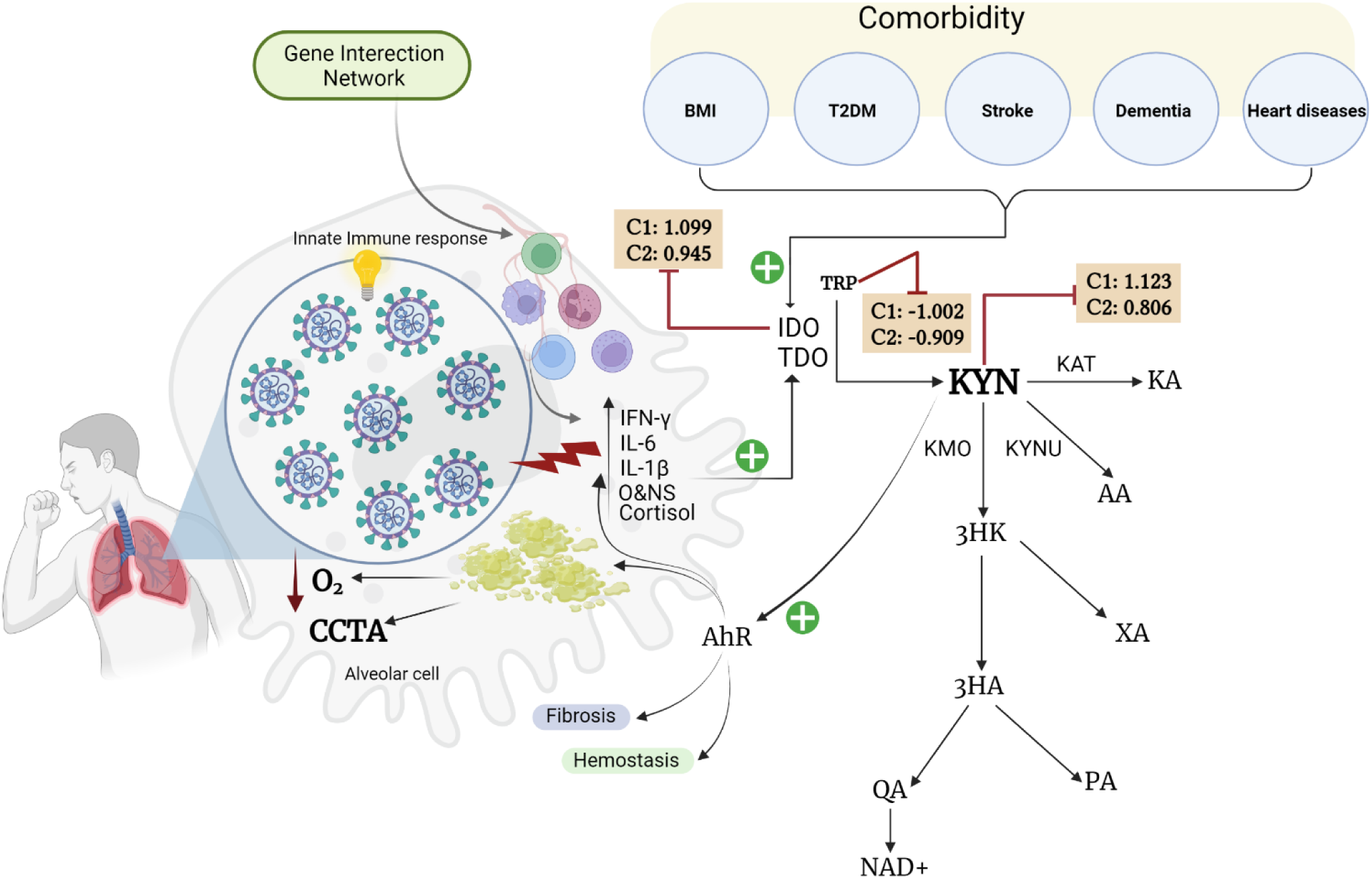
Summary of TRYCAT pathway in COVID-19. BMI: Body Mass Index, C: Cohort, T2DM: Type 2 Diabetes Mellitus, IFN-γ: Interferon-Gamma, IL-6: Interleukin 6, IL-1β: Interleukin-1 beta, O&NS: Oxidative and nitrosative stress, O_2_: Oxygen, CCTA: Chest Computed Tomography Abnormalities, AhR: Aryl Hydrocarbon Receptor, IDO: Indoleamine 2,3 dioxygenase, TDO: Tryptophan 2,3-dioxygenase, KAT: Kynurenine Aminotransferase, KMO: Kynurenine 3-monooxygenase, KYNU: Kynureninase, TRP: Tryptophan, KYN: Kynurenine, KA: Kynurenic Acid, 3HK: 3-Hydroxykynurenine, AA: Anthranilic Acid, XA: Xanthurenic Acid, 3HA: 3-Hydroxyanthranilic Acid, PA: Picolinic Acid, QA: Quinolinic Acid, NAD^+^: Nicotinamide adenine dinucleotide.

Nevertheless, no systematic review and meta-analysis were conducted in COVID-19 and severe/critical COVID-19 to examine whether the TRYCAT pathway is activated. Hence, the purpose of the current study was to systematically review and meta analyze the TRP and TRYCAT results in COVID-19 patients versus controls and severe/critical versus mild/moderate COVID-19.

## Material and Methods

The current meta-analysis was in compliance with the standards of Preferred Reporting Items for Systematic Reviews and Meta-Analyses (PRISMA) 2020 (Page, McKenzie et al. 2021), the guidelines of the Cochrane Handbook for Systematic Reviews and Interventions (Higgins JPT 2019), along with the Meta-Analyses of Observational Studies in Epidemiology (MOOSE). In the present meta-analysis, we examine TRP and TRYCATs levels. We also compute indices of IDO enzyme activity namely the KYN/TRP and (KA+KYN)/TRP ratios, and a proxy of KAT enzyme activity namely the KA/KYN and KA/(KYN+TRP) ratios.

### Search strategy

The search started December 15,2021 and at the end of this month all required data were collected. We entered specific mesh terms and keywords in electronic databases to find related articles in PubMed/MEDLINE, Google Scholar, and Web of Science. These terms and keywords, focused on TRP, TRYCATs and COVID-19, are shown in the Electronic Supplementary File (ESF), Table 1. To ensure that we included all the related articles, we also searched the reference list of previous reviews and grey literature.

**Table 1.** The number of COVID-19 patients and studies included in the meta-analyses and the side of standardized mean difference (SMD) and the 95% confidence intervals with respect to the zero SMD.

| Outcome profiles | n studies | Side of 95% confidence intervals |  |  |  | Patient | Control | Total |
| --- | --- | --- | --- | --- | --- | --- | --- | --- |
|  |  | < 0 | Overlap 0<br>and SMD < 0 | Overlap 0<br>and SMD > 0 | > 0 | Cases | Cases | number of<br>participants |
| Cohort 1: COVID-19 Patients Versus Non-COVID-19 Control |  |  |  |  |  |  |  |  |
| KYN/TRP | 10 | 1 | 0 | 2 | 7 | 329 | 475 | 804 |
| KYN | 8 | 0 | 1 | 0 | 7 | 285 | 419 | 704 |
| TRP | 7 | 5 | 1 | 1 | 0 | 275 | 409 | 684 |
| KA | 4 | 0 | 1 | 2 | 1 | 113 | 94 | 207 |
| KA/(KYN+TRP) | 8 | 1 | 1 | 4 | 2 | 285 | 419 | 704 |
| (KYN+KA)/TRP | 10 | 1 | 0 | 3 | 6 | 329 | 475 | 804 |
| KA/KYN | 8 | 3 | 3 | 2 | 0 | 285 | 419 | 704 |
| Cohort 2: Severe/Critical COVID-19 patients Versus Mild/Moderate COVID-19 patients |  |  |  |  |  |  |  |  |
| KYN/TRP | 9 | 0 | 0 | 1 | 8 | 270 | 503 | 773 |
| KYN | 6 | 0 | 0 | 3 | 3 | 184 | 399 | 583 |
| TRP | 5 | 3 | 2 | 0 | 0 | 153 | 282 | 435 |

KYN: kynurenine, TRP: tryptophan, KA: kynurenic acid

### Eligibility standards

We included published papers in peer-reviewed journals and the English language as the main criteria for selecting articles. However, we also reviewed manuscripts published in other languages such as Thai, French, Spanish, German, Italian, Arabic. Inclusion criteria were: a) observational case-control and cohort studies which assessed quantitatively the concentration of TRP and TRYCATs in serum, plasma, CSF, and brain tissues of patients who showed a positive real-time polymerase chain reaction (RT-PCR test and were either symptomatic or asymptomatic; b) studies reporting data in a control group comprising healthy people, previously infected or recovered patients, or a subgroup of mild/moderate COVID-19 patients; and c) the results are reported as quantitative scores with mean and standard deviation (SD) or standard error of the mean (SEM). We excluded the following studies: a) systematic and narrative reviews and meta-analysis studies; b) duplicate studies as well as animal and genetic studies; c) articles that used other media including saliva; and d) the authors did not show mean and SD/SEM of the measured biomarkers or any other mean to estimate these values. When the authors presented geometric means, median (interquartile range, range), or represented data as a graph, we sent emails to request the mean ±SD in the study groups. Without response of the authors, we used the estimation method described by Wan and Wang (Wan, Wang et al. 2014) to compute mean ±SD from median with either interquartile range or range. In addition, Web Plot Digitizer was employed to extract quantitative data represented in a graph (https://automeris.io/WebPlotDigitizer/).

### Primary and secondary outcomes

The primary outcome is IDO activity which we assessed through the KYN/TRP and (KYN+KA)/TRP ratios (Bonaccorso, Marino et al. 2002) in COVID-19 patients versus controls. Secondary outcomes were the KA/KYN and KA/(KYN+TRP) indices which reflect KAT enzyme activity, while the KA/KYN ratio also reflects neurotoxicity. If significant, we also examined the separate TRP, KYN, and KA results. The TRP and TRYCATs data were not only examined in COVID-19 versus non-COVID-19 controls (study cohort 1) but also in severe/critical (including death due to COVID-19) versus mild/moderate COVID-19 (study cohort 2).

### Screening and data extraction

The first author (AA) performed an initiatory review by evaluating the titles and abstracts to ensure which papers were eligible to be included. Consequently, eligible full-text articles were downloaded after removing some publications according to the predetermined exclusion criteria. All required data extracted from the articles were entered in a predefined excel spreadsheet file made for this project, including researcher’s names, publication date, quantitative data of TRP and TRYCATs, the number of the participants either as a COVID-19 or control groups, demographic data such as age (expressed as mean ±SD), male/female count, type of sample, serum or plasma, severity level, country latitude in which the study was conducted, and quality scores of the studies (see below). Furthermore, all extracted data in the excel spreadsheet were scrutinized by the second author (TS) immediately after the first author finalized entering the data. The last author (MM) was consulted in the case of controversial results. The last author slightly adjusted the “immune confounder scale (ICS)” published previously (Andrés-Rodríguez, Borràs et al. 2019) to estimate the methodological quality of TRYCATs studies. This ICS and the related repoint checklist are shown in ESF, Table 2.

**Table 2.** Results of meta-analysis performed on several outcome variables of the tryptophan catabolite (TRYCAT) pathway

| Outcome feature sets | n | Groups | SMD | 95% CI | z | p | Q | df | p | I<sup>2</sup> (%) | $\tau^2$ | T |
| --- | --- | --- | --- | --- | --- | --- | --- | --- | --- | --- | --- | --- |
| ***Cohort 1: COVID-19 Patients Versus Non-COVID-19 Control*** |  |  |  |  |  |  |  |  |  |  |  |  |
| KYN/TRP | 10 | Overall | 1.099 | 0.714; 1.484 | 5.594 | 0.000 | 85.310 | 9 | 0.000 | 89.450 | 0.616 | 0.785 |
|  | 4 | Serum | 1.359 | 0.889;1.829 | 5.666 | 0.000 | 9.104 | 3 | 0.028 | 67.049 | 0.145 | 0.390 |
|  | 6 | Plasma | 0.569 | -0.103;1.241 | 1.661 | 0.097 | 38.093 | 5 | 0.000 | 86.874 | 0.591 | 0.769 |
|  | Q= 3.568, df=1, p=0.059 |  |  |  |  |  |  |  |  |  |  |  |
| KYN | 7 | Overall | 1.123 | 0.730;1.516 | 5.595 | 0.000 | 29.745 | 7 | 0.000 | 76.467 | 0.221 | 0.470 |
| TRP | 7 | Overall | -1.002 | -1.738; -0.266 | -2.668 | 0.008 | 88.926 | 6 | 0.000 | 93.253 | 0.892 | 0.944 |
| KA | 4 | Overall | 0.164 | -0.120;0.449 | 1.133 | 0.257 | 6.947 | 3 | 0.074 | 56.816 | 0.128 | 0.358 |
|  | 2 | Serum | 0.649 | 0.170;1.129 | 2.653 | 0.008 | 0.617 | 1 | 0.432 | 0.000 | 0.000 | 0.000 |
|  | 2 | Plasma | -0.098 | -0.451;0.255 | -0.546 | 0.585 | 0.275 | 1 | 0.600 | 0.000 | 0.000 | 0.000 |
|  | Q = 6.055, df=1, p=0.014 |  |  |  |  |  |  |  |  |  |  |  |
| KA/(KYN+TRP) | 10 | Overall | 0.297 | 0.089;0.506 | 2.794 | 0.005 | 40.957 | 9 | 0.000 | 78.026 | 0.255 | 0.505 |
| (KYN+KA)/TRP | 10 | Overall | 0.789 | 0.261;1.318 | 2.926 | 0.003 | 87.188 | 9 | 0.000 | 89.678 | 0.624 | 0.790 |
| KA/KYN | 8 | Overall | -0.398 | -0.967; 0.170 | -1.373 | 0.170 | 66.867 | 7 | 0.000 | 89.531 | 0.569 | 0.754 |

| *Cohort 2: Severe/Critical COVID-19 patients Versus Mild/Moderate COVID-19 patients* |  |  |  |  |  |  |  |  |  |  |  |  |
| --- | --- | --- | --- | --- | --- | --- | --- | --- | --- | --- | --- | --- |
| KYN/TRP | 8 | Overall | 0.945 | 0.629; 1.262 | 5.848 | 0.000 | 25.776 | 8 | 0.001 | 68.964 | 0.151 | 0.389 |
| KYN | 6 | Overall | 0.806 | 0.462; 1.149 | 4.593 | 0.000 | 13.343 | 5 | 0.020 | 62.528 | 0.108 | 0.328 |
| TRP | 5 | Overall | -0.909 | -1.569; -0.249 | -2.699 | 0.007 | 28.165 | 4 | 0.000 | 85.798 | 0.462 | 0.680 |

KYN: kynurenine, TRP: tryptophan, KA: kynurenic acid

These scores estimate key quality data such as sample size, covariate control and the time of sampling. The best methodological quality is obtained when the ICS score is close to 10 with the overall score ranging from 0 to 10. The redpoints score scale mainly focuses on the poor adjustment of the key confounders, which may cause biased TRYCATs results (either due to biological or analytical variation), along with an uncontrolled study design. The range of the score scale is from 0 to 26 with values close to 26 indicating poor control and quality.

### Data analysis

We employed the CMA V3 software to conduct the current meta-analysis and we followed the PRISMA guidelines (Page, McKenzie et al. 2021). The presence of TRYCATs in at least three studies was the determinant for conducting a meta-analysis. The biomarker’s outcomes as assessed in our systematic review and meta-analysis are displayed in **Table 1**. By calculating the mean values of the markers in their respective profiles (e.g. KYN/TRP ratio) and assuming dependency, we compared the synthetic scores indicating these profiles in COVID-19 patients (or subgroups) versus their controls. In the meta-analysis, IDO activity was estimated by specifying the direction of the effect size of KYN as positive (favoring COVID-19) and TRP as negative. Moreover, KAT activity namely the KA/(KYN+TRP) and KA/KYN ratios, was estimated by entering KA with positive direction (thus favoring COVID-19), and TRP and/or KYN with negative direction in the meta-analysis. A restricted maximum-likelihood random-effects model was utilized based on our hypothesis that the included studies have different characteristics. The standardized mean difference (SMD) with 95% confidence intervals (95% CI) was computed as the indicator for the effect size. We considered the results to be statistically significant when p <0.05 (two-tailed tests). SMD values of 0.8, 0.5, and 0.2 indicate large, moderate, and small effect sizes, respectively (Cohen 2013). Heterogeneity was examined by tau-squared values as mentioned previously and we also computed the Q and I^2^ metrics (Vasupanrajit, Jirakran et al. 2021, Vasupanrajit, Jirakran et al. 2021). We also used the leave-one-out approach to conduct sensitivity analyses to assess the robustness of the pooled combined meta-analysis effects and between-study heterogeneity. We assessed possible differences in TRP and TRYCATs between serum and plasma (Almulla, Vasupanrajit et al. 2021) by considering these subgroups as a unit of analysis. We compared the effects at different study levels and ran the meta-analysis across subgroups. We assessed the impact of small study effects, including publication bias, using the conventional fail-safe N approach, Kendall tau with continuity correction (using one-tailed p-values), and Egger’s regression intercept (using one tailed p-values). When Egger’s linear regression test indicates substantial asymmetry, we estimate the modified effect size after accounting for the impacts of missing studies using Duval and Tweedie’s trim-and-fill approach.

## Results

### Search findings

During the selection process, 30 articles were investigated in the current study based on the keywords shown in ESF, table 1. The detailed information related to the inclusion-exclusion criteria of the research papers and the outcomes of our search process are presented in the PRISMA flowchart shown in **Figure 2**. Nineteen full-text papers were eligible for the systematic review after 11 records were removed from the initial number of articles. Finally, the meta-analysis involved 14 articles as 5 papers were excluded for reasons listed in ESF, Table 3 (Blasco, Bessy et al. 2020, Fraser, Slessarev et al. 2020, Kimhofer, Lodge et al. 2020, Robertson, Gostner et al. 2020, Thomas, Stefanoni et al. 2020, Ansone, Briviba et al. 2021, D’Amora, Silva et al. 2021, Herrera-Van Oostdam, Castaneda-Delgado et al. 2021, Lawler, Gray et al. 2021, Lionetto, Ulivieri et al. 2021, Mangge, Herrmann et al. 2021, Marin-Corral, Rodriguez-Morato et al. 2021, Michaelis, Zelzer et al. 2021, Xiao, Nie et al. 2021).

**Figure 2:**
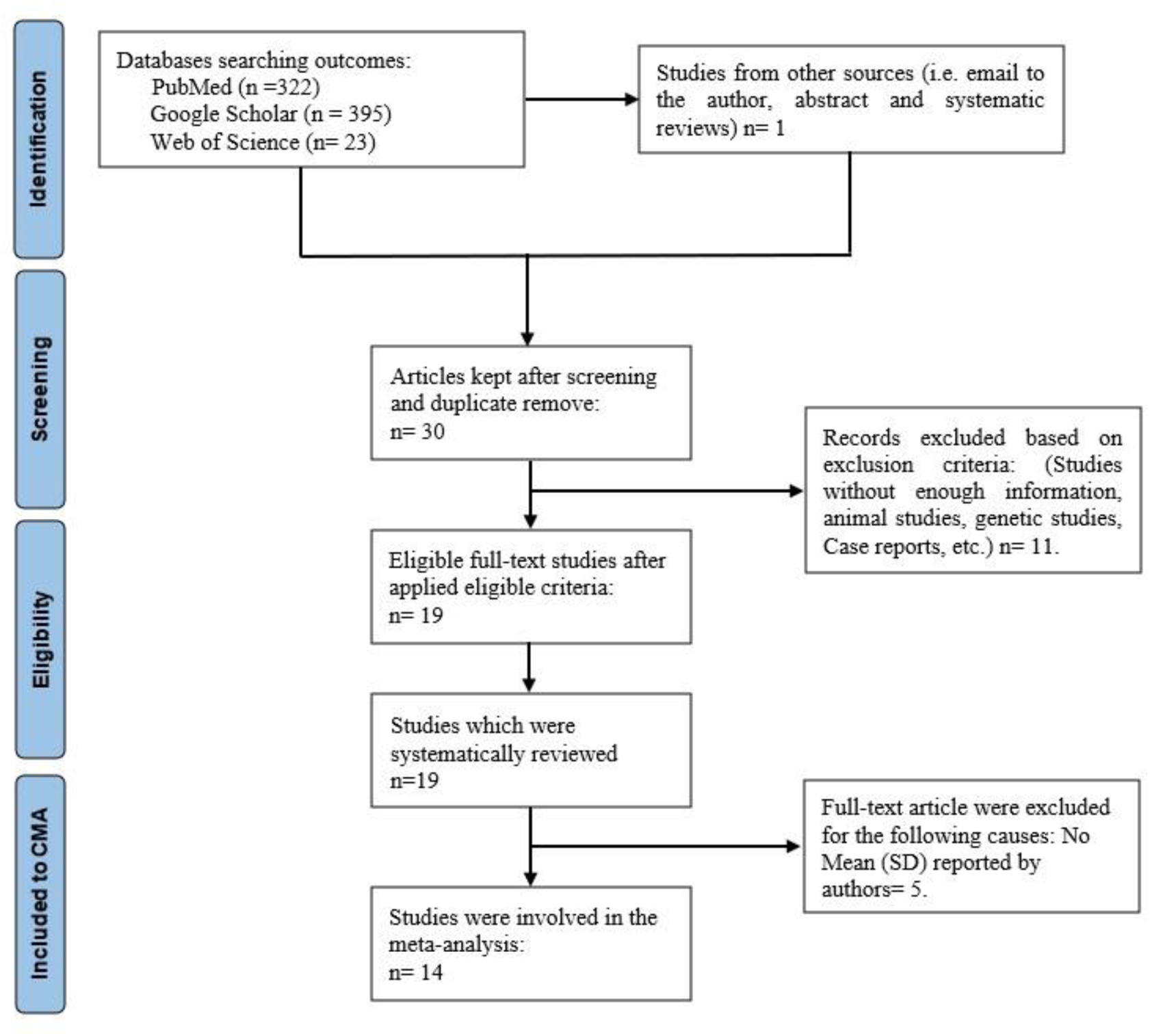
Prisma Flow Chart

**Table 3.** Results on publication bias.

| Outcome feature sets | Fail safe n | Z Kendall's $\tau$ | p | Egger's t test (df) | p | Missing studies (side) | After Adjusting |
| --- | --- | --- | --- | --- | --- | --- | --- |
| *Cohort 1: COVID-19 Patients Versus Non-COVID-19 Control* |  |  |  |  |  |  |  |
| KYN/TRP (Overall) | 10.499 | 0.089 | 0.464 | 0.989 (8) | 0.175 | 3 (left) | SMD= 0.573 0.003; 1.144) |
| KYN/TRP (Serum) | 11.244 | 0.679 | 0.248 | 1.223 (2) | 0.172 | 0 |  |
| KYN/TRP (Plasma) | 4.373 | 0.563 | 0.286 | 0.347 (4) | 0.372 | 1 (left) | SMD= 0.462 (-0.155; 1.080) |
| KYN (Overall) | 11.647 | 0.494 | 0.310 | 0.269 (6) | 0.398 | 2 (left) | SMD= 0.961 (0.584; 1.338) |
| TRP (Overall) | -11.267 | 0.150 | 0.440 | 0.947 (5) | 0.193 | 1 (Right) | SMD= -0.817 (-1.618; -0.017) |
| KA (Overall) | 2.478 | 0.000 | 0.500 | 0.720 (2) | 0.273 | 0 |  |
| KA/(KYN+TRP)<br>(Overall) | 1.673 | 1.162 | 0.122 | 0.549 (8) | 0.298 | 1 (Left) | SMD=0.023 (-<br>0.358;0.405) |
| (KYN+KA)/TRP<br>(Overall) | 9.818 | 0.804 | 0.210 | 1.213 (8) | 0.129 | 2 (Left) | SMD= 0.579 0.0007;<br>1.157) |
| KA/KYN<br>(Overall) | -5.376 | 0.247 | 0.402 | 1.598 (6) | 0.080 | 0 |  |
| ***Cohort 2: Severe/Critical COVID-19 patients Versus Mild/Moderate COVID-19 patients*** |  |  |  |  |  |  |  |
| KYN/TRP<br>(Overall) | 10.226 | 1.251 | 0.105 | 2.362 (7) | 0.025 | 1 (Left) | SMD=0.876 (0.562;<br>1.189) |
| KYN<br>(Overall) | 7.758 | 0.187 | 0.425 | 0.129 (4) | 0.451 | 0 |  |
| TRP<br>(Overall) | -5.887 | 1.959 | 0.025 | 5.897 (3) | 0.004 | 0 |  |

In this meta-analysis, we considered case-control and retrospective studies. This study examined 804 subjects in study cohort 1 (329 COVID-19 patients and 475 non-COVID-19 controls), and 773 individuals in the second cohort (involving 270 severe/critical COVID-19 patients and 503 mild/moderate COVID-19 patients). As shown in ESF, table 3, we excluded 5 studies from the meta-analysis. Cohort 1 examined TRYCATs in plasma in 6 studies, while 4 studies used serum, whereas in the second cohort, 6 studies were based on plasma and 3 used serum. Liquid chromatography-mass spectrometry (LC-MS) was used in 5 studies, while 3 studies utilized liquid chromatography and two mass spectrometry (LC-MS/MS). Ultra-high-performance liquid-chromatography-mass spectrometry (UHPLC-MS) and high-performance liquid chromatography were employed in two studies and the remaining studies used liquid chromatography–high-resolution mass spectrometry (LC-HRMS) and liquid chromatography-UV detection (LC-UV). All the included studies were conducted on patients who showed positive COVID-19 with different severity. In the control group, some authors reported that they never got an infection with covid-19 and others just mentioned they were not infected.

Overall, within 14 eligible studies, there were 794 covid-19 patients (329 in case control studies and 465 in retrospective studies) and 475 non-covid-19 controls. Some of the patients from case control studies were included in both cohorts of the meta-analysis in case the author reported on severe/critical and mild/moderate COVI-19 and control groups. The ages of the participant were between 40 and 95 years old. Brazil, USA, Latvia, Canada, France, China, Mexico, Sweden, Spain, and Italy contributed with 1 study, and Australia and Austria with 2 studies. However, most participants were from Italy due to the large sample size. ESF, Table 4 shows the median (min-max) of ICS namely redpoint and quality scores which equaled 12.5 (min= 6, max=17) and 3 (min=3, max=7), respectively.

## COVID-19 versus controls

### The primary outcome variables KYN/TRP and (KYN+KA)/TRP ratio

The results of the systematic review on KYN/TRP in COVID-19 are shown in Table 1. We found that out of the ten included studies, the 95% CI for 7 (4 serum, 3 plasma) were entirely on the positive side of zero, while only one (plasma) study was totally on the negative side of zero. The two other studies showed 95% CI intervals which overlapped with zero but with SMD values that were greater than zero. **Figure 3** shows the forest plot of KYN/TRP in COVID-19 patients versus non-COVID-19 controls. We performed subgroup analyses to examine the high heterogeneity as indicated by elevated values of τ^2^. These results showed a trend toward a possible difference (p=0.059) between serum and plasma. The serum results displayed a huge and significant effect size between COVID-19 and controls, whereas the plasma findings were non-significant (**Table 2**). In addition, in serum, the heterogeneity was lower as compared with plasma. We found 3 missing studies on the left side and imputation of these missing studies lowered the SMD to 0.573 (95% CI: 0.003; 1.144), although still significant. Serum results did not show any bias; while there was 1 missing study in plasma and after imputation the SMD decreased to 0.462 (95% CI: -0.155; 1.080).

**Figure 3:**
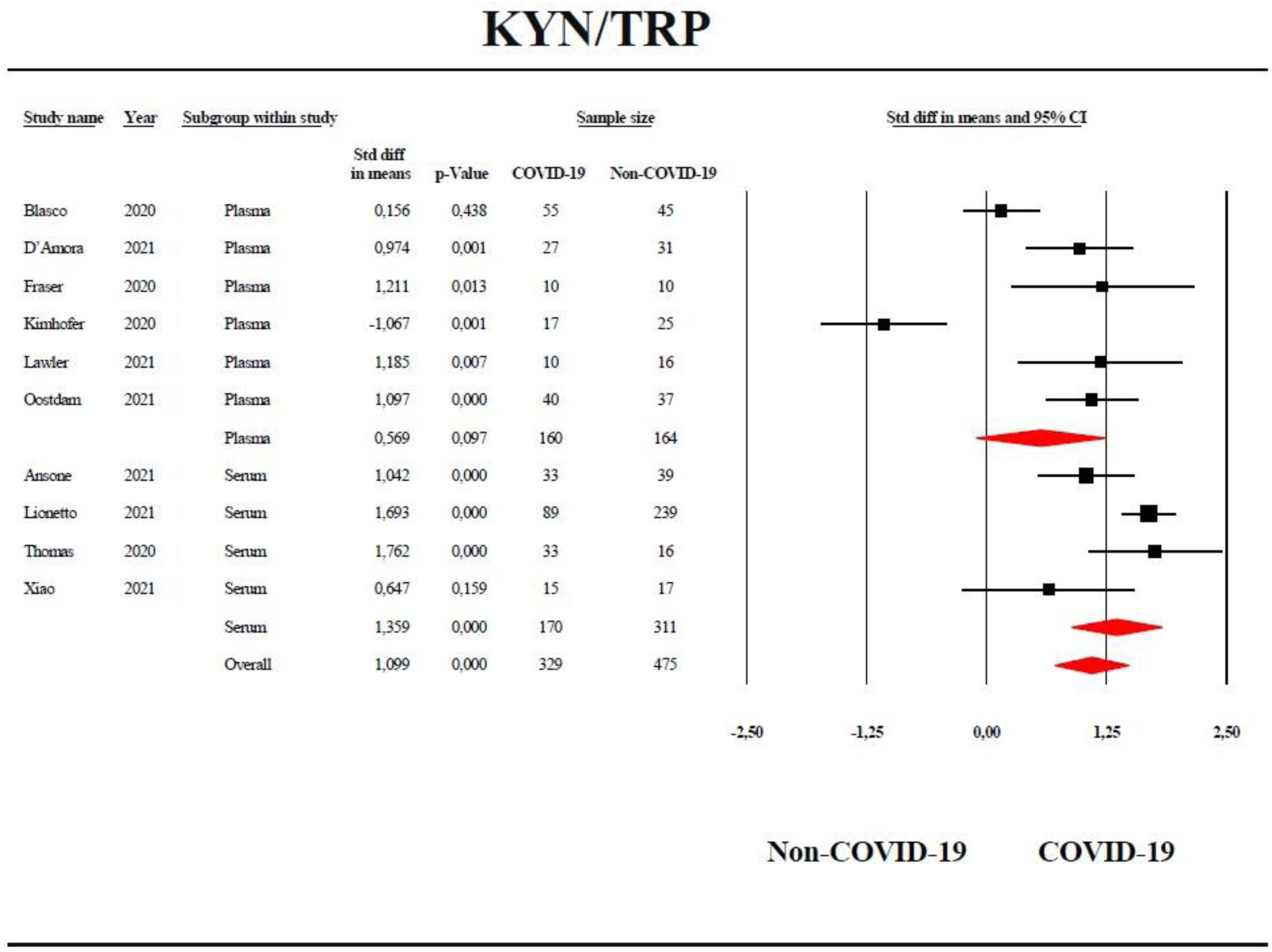
Forest plot with the results of a meta-analysis performed on the kynurenine / tryptophan (KYN/TRP) ratio in COVID-19 patients versus non-COVID-19 controls.

Table 1 shows that in 6 studies (out of 10) the 95% CI were totally on the right side of zero and that only one study showed that the CI were completely on the left side of zero. The 95% CI of the other 3 studies crossed the zero line but all showed SMD values that were greater than zero. Table 2 shows there is a significant difference in (KYN+KA)/TRP ratio between COVID-19 patients and controls with a large effect size of SMD=0.789. ESF, Figure 1 displays the forest plot of the (KYN+KA)/TRP ratio. **Table 3** shows no evidence of publication bias although there were two missing studies on the left side and a somewhat lower adjusted SMD after imputation (SMD=0.579).

Table 1 and ESF, Figure 2 display the forest plot of TRP in COVID-19. Table 2 shows an overall significant decrease in TRP in COVID-19 with a high effect size (SMD=- 1.002). Although there were no significant differences (p=0.404) between plasma and serum, serum TRP was significantly decreased in COVID-19 (SMD=-1.216), whereas plasma TRP did not show significant differences. Table 3 shows no evidence of publication bias although Duval and Tweedie’s trim and fill showed one missing value on the right side and imputation yielded an adjusted SMD of -0.817.

Table 1 shows that out of 8 KYN studies, the 95% CI of 7 studies was completely on the right side of zero, while one CI intersected with zero. ESF, Figure 3 and Table 2 show the KYN results indicating a highly significant increase in KYN in COVID-19 (SMD= 1.123). Duval and Tweedie’s trim and fill showed two missing studies on the left side and imputing these studies yielded a slightly decreased effect size (SMD= 0.961, 95% CI: 0.584; 1.338), which was still significant.

### Secondary outcome variables

#### KA/KYN ratio

Table 1 and ESF, Figure 4 show the KA/KYN ratio in 8 studies, of which 3 showed 95% CI that were completely on the left side of zero and 5 studies that overlapped with zero. Table 2 shows that there were no significant differences in the KA/KYN ratio between COVID-19 and controls. The KA/KYN results were free from publication bias.

**Figure 4:**
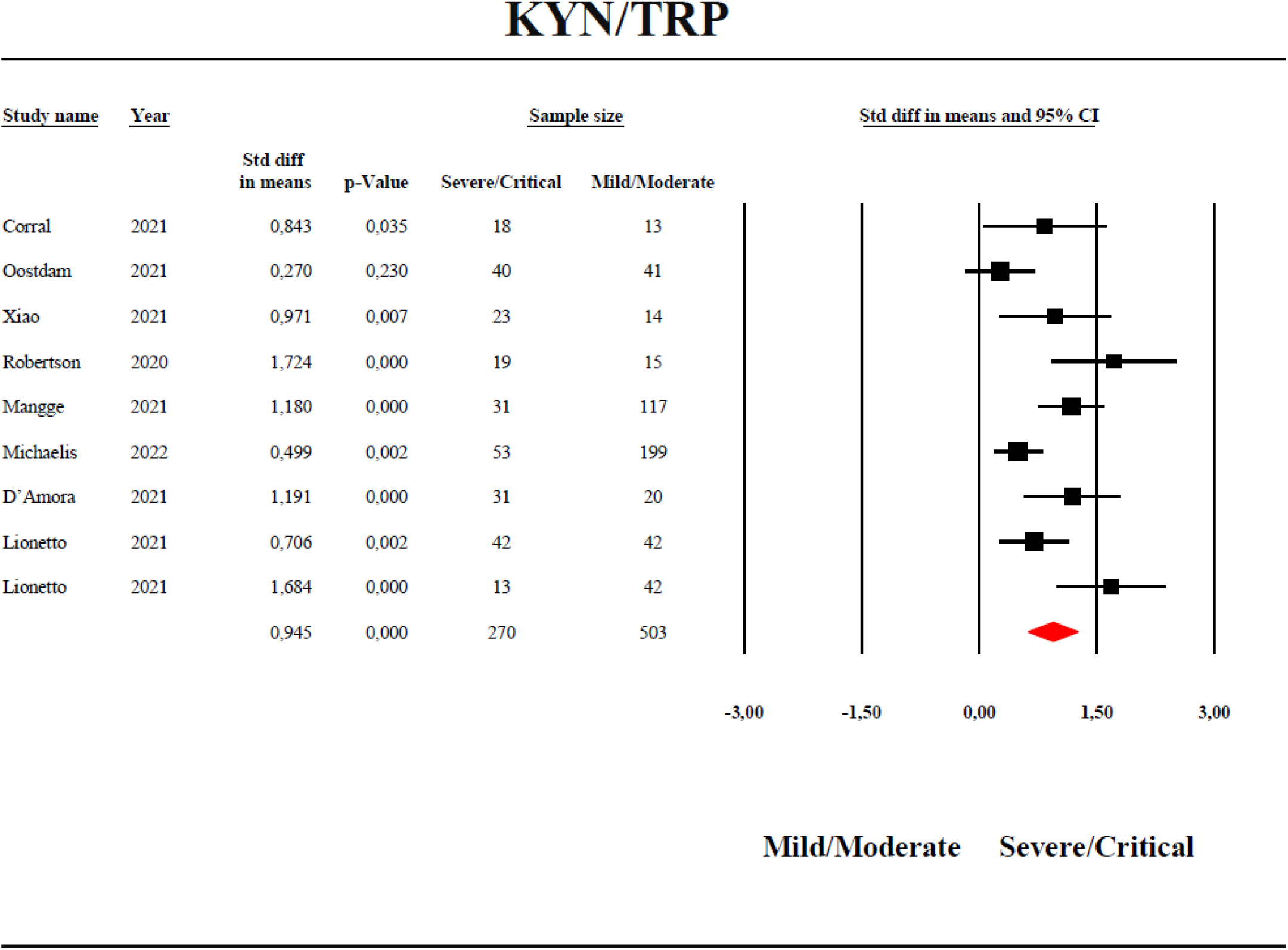
Forest plot with the results of a meta-analysis performed on the kynurenine / tryptophan (KYN/TRP) ratio in severe/critical COVID-19 versus mild/moderate COVID-19.

#### KA/(KYN+TRP) ratio

Table 1 and ESF, Figure 5 show that the KA/(KYN+TRP) results in 10 studies and ESF, Figure 5 shows the forest plot. There was a significant increase with a small effect size (SMD= 0.297) in KA/(KYN+TRP) in COVID-19 patients as compared with controls. However, after imputing one missing study, the SMD decreased to 0.023 and was no longer significant (Table 3).

#### KA levels

KA results were obtained in 4 studies. Table 1 and ESF Figure 6 shows that 3 studies intersected with zero with 2 studies showing SMD values greater than zero and one study less than zero, while one study showed 95% CI which were completely on the right side of zero. There was a high heterogeneity when serum and plasma were combined with a significant difference (p=0.014) between both media. Therefore, we conducted a subgroup analyses showing that the results in serum contradicted those in plasma (see Table 2).

#### Severe/critical COVID-19 versus mild/moderate COVID-19

Table 1 shows that all 95% CI of the cohort 2 studies reporting on severe/critical versus mild/moderate COVID-19 were completely on the right side of zero (favoring severe/critical patients), except 1 study which crossed zero. There is a significant difference in the KYN/TRP ratio between severe/critical versus mild/moderate COVID-19 with a huge effect size favoring severe/critical COVID-19 (SMD= 0.945). **Figure 4** shows the forest plot of the KYN/TRP ratio between severe/critical versus mild/moderate patients. Publication bias with one missing study was detected and the adjusted SMD was slightly less than the observed but was still significant with a high impact size (SMD=0.876). ESF, Figure 7 and Table 2 show significantly lowered TRP in severe/critical COVID-19 as compared with mild/moderate COVID-19 with a large effect size (SMD= -0.909). Table 2 and ESF, Figure 8 show an overall significant difference in KYN level between severe/critical and mild/moderate COVID-19 with a large effect size (SMD=0.806). Table 3 did not show evidence of publication bias in the KYN data in critical COVID-19.

#### Meta-regression analysis

The meta-regression results are shown in ESF, table 5 indicating that the type of medium significantly affected the KYN/TRP (p=0.047) and (KYN+KA)/TRP (p=0.051) ratios, and also KA (p=0.006). Furthermore, the sample size had a significant effect (p=0.022) on the KA/KYN ratio, and the severity of disease exerted a significant effect (p= 0.003) on the KA/(KYN+TRP) ratio.

## Discussion

The first major findings of this systematic review and meta-analysis are that a) the KYN/TRP ratio is significantly increased in COVID-19 patients compared to non-COVID-19 controls with high effect size; and b) the KYN/TRP ratio is dramatically increased in severe/critical COVID-19 as compared with mild/moderate COVID-19 again with a large effect size. Importantly, the severe/critical COVID-19 patient samples included in this study mainly consist of critical patients who did not survive and, therefore, our results also suggest that an increased KYN/TRP ratio is associated with death due to COVID-19.

These results indicate that IDO activity and the TRYCAT pathway are upregulated in COVID-19 and that it predicts critical disease and non-survival. The most probable cause of IDO enzyme activation in COVID-19 is the increased level of pro-inflammatory cytokines including IFN-γ, IL-1β and IL-6 (Akbari, Tabrizi et al. 2020, Mulchandani, Lyngdoh et al. 2021) and activated oxidative stress pathways (Doğan, Bal et al. 2021), which both potently stimulate IDO (Smith and Maes 1995, Lionetto, Ulivieri et al. 2021).

Further analyses showed that the changes in the KYN/TRP ratio are attributable to significant increases in KYN and decreases in TRP in COVID-19 again with large effect sizes. These results extend the findings of previous studies which showed associations between severity of COVID-19 and increases in the KYN/TRP ratio and KYN and decreases in TRP (Lionetto, Ulivieri et al. 2021, Marin-Corral, Rodriguez-Morato et al. 2021, Michaelis, Zelzer et al. 2021). Thus, not only aberrations in innate immune potential but also associated TRYCAT pathway activation contributes to a fatal course of the disease (Blanco-Melo, Nilsson-Payant et al. 2020, Ragab, Salah Eldin et al. 2020, Ombrello and Schulert 2021).

The second major finding of this study is that the KA/KYN ratio did not show a significant difference between COVID-19 patients as compared to non-COVID-19 controls, suggesting that COVID-19 is not accompanied by changes in KAT activity. Our meta-analysis performed on KA values in COVID-19 showed important heterogeneity and subsequent groups analysis revealed that serum KA was significantly increased in COVID-19 with medium effect size (0.649), whereas in plasma a non-significant inverse association was found. There are insufficient data to perform meta-analysis on other ratios reflecting KMO and KYNU activity. In this respect, Lawler et al., reported elevated levels of 3HK and QA in patients with COVID-19 compared to healthy controls (Lawler, Gray et al. 2021). Likewise, Marin-Corral et al. reported a high level of 3HK in severe/critical COVID-19 patients compared to those with mild/moderate infection (Marin-Corral, Rodriguez-Morato et al. 2021).

During infection, IDO activation and consequent increased TRYCATs but lowered TRP levels are key components of the innate immune response. First, the TRYCAT pathway has major intrinsic scavenging activities by neutralizing ROS (Maes, Leonard et al. 2011). Moreover, some TRYCATs have antioxidant properties on their own as for example, 3-hydroxyanthranilic acid (3HA) and 3HK, which are more effective as radical scavengers than tocopherol, and XA, which has antioxidant activity comparable to that of butylated hydroxytoluene (BHT) (Goda, Hamane et al. 1999, Maes, Leonard et al. 2011). KA has adequate antioxidant effects by protecting tissues from oxidative damage (Lugo-Huitrón, Blanco-Ayala et al. 2011, Pérez-González, Alvarez-Idaboy et al. 2015). Second, reduced TRP exerts anti-inflammatory (reduced T cell proliferation and activation, sensitization of apoptosis of activated T cells, and induction of the regulatory phenotype) and antimicrobial (inhibiting the growth of virus, bacteria and parasites) effects through TRP starvation (Mellor and Munn 1999, Lee, Park et al. 2002, Fallarino, Grohmann et al. 2006, Schmidt, Muller et al. 2009, Yan, Zhang et al. 2010). Third, TRYCATs such as KA, KYN, QA, and XA, may exert negative immune-regulatory effects by lowering the production of IFN-γ and/or increasing that of IL-10 (Maes, Mihaylova et al. 2007, Maes, Leonard et al. 2011). In addition, KA has potent anti-inflammatory effects while diminished KA levels may aggravate tissue damage and cell proliferation (Wirthgen, Hoeflich et al. 2018). IFN-γ-induced stimulation of antigen-presenting cells upregulates the TRYCAT pathway and results in a counter-regulatory effect that preserves homeostasis (Mandi and Vecsei 2012). Due to the fact that TRYCATs trigger apoptosis in Th-1, but not Th-2, cells, TRYCAT pathway activation may suppress Th-1 cells but promotes Th-2 cell survival (Fallarino, Grohmann et al. 2002, Xu, Zhang et al. 2008). As such, TRYCAT pathway activation results in a negative feedback loop to limit ROS production, hyperinflammation, and the Th-1 response (Maes, Mihaylova et al. 2007, Xu, Zhang et al. 2008). Fourth, some TRYCATs have neuroprotective effects including KA, anthranilic acid (AA) and XA. Thus, KA may inhibit N-methyl-D-aspartate (NMDA), kainate glutamate ionotropic, and amino-3-hydroxy-5-methyl-4-isoxazolepropionic acid (AMPA) receptors, and reduce glutamate liberation through attenuating alpha 7 nicotinic acetylcholine receptors (Morris, Carvalho et al. 2016, Almulla and Maes 2022). XA inhibits vesicular glutamate transport (VGLUT), synaptic transmission via the NMDAR receptor, and excitatory postsynaptic potentials (Kanchanatawan, Sirivichayakul et al. 2018). Furthermore, AA has neuroprotective effects by blocking the synthesis of neurotoxic TRYCATs such picolinic acid (PA) and QA from 3HA (Guillemin, Cullen et al. 2007). Based on the above, it appears that during infection, the TRYCAT pathway activation has major homeostatic effects.

Nevertheless, overproduction of some TRYCATs may cause detrimental effects in COVID-19. KA is implicated in deteriorating male COVID-19 patients through affecting the AhR, one of the master regulators of the immune-inflammatory response (Cai, Kim et al. 2021). In addition, activation of AhR by TRYCATs, mainly KYN, affects immune resistance against viral infections and the airway basal cells of the lung epithelium, which are responsible for tissue repair (Giovannoni, Li et al. 2020, Thomas, Stefanoni et al. 2020). Most importantly, corona viruses activate the same receptor through an IDO-independent mechanism while the IDO-AhR pathway in employed by viruses, bacteria, and parasites to establish infection (Turski, Wnorowski et al. 2020). Consequently, a positive feedback loop is established between increased TRYCATs levels due to IDO activation and stimulation of the AhR by TRYCATs and corona virus (Turski, Wnorowski et al. 2020). Moreover, the AhR may enhance IDO transcription and regulates IDO activity (Pallotta, Fallarino et al. 2014). These processes may result in the SAAS which may result in activated immune-inflammatory pathways (increased M1 cytokines), fibrosis (increased IL-22), thromboembolism (increased tissue factor and plasminogen activator inhibitor 1), consequent multiple organ injuries including brain injuries, and eventually death (Turski, Wnorowski et al. 2020).

Moreover, some TRYCATs have depressogenic, anxiogenic and neurotoxic effects and TRYCATs like KYN are increased in neuropsychiatric illness including major depression, anxiety, and psychosis (Maes, Leonard et al. 2011, Anderson, Maes et al. 2013). Second, some TRYCATs exhibit pro-oxidant properties as evidenced by increased ROS, hydrogen peroxide, and superoxide production, and increased oxidative damage including lipid peroxidation caused by 3HA, 3HK, and QA (Dykens, Sullivan et al. 1987, Okuda, Nishiyama et al. 1998, Guidetti and Schwarcz 1999, Goldstein, Leopold et al. 2000, Santamaría, Galván-Arzate et al. 2001, Smith, Smith et al. 2009, Reyes Ocampo, Lugo Huitrón et al. 2014). Third, TRYCATs such as QA and XA and PA may have direct neurotoxic effects by activating hippocampal NMDAR and causing excitotoxicity with apoptosis and hippocampal shrinkage thereby inducing neurocognitive impairments (Lugo-Huitron, Ugalde Muniz et al. 2013, Rahman, Rao et al. 2018). Elevated XA levels may cause severe neuronal damage, apoptosis, mitochondrial dysfunctions, disrupt glutamate transmission, and impair presynaptic transmission caused by NMDAR stimulation (Kanchanatawan, Sirivichayakul et al. 2018). Such effects may contribute to the development of neuropsychiatric disorders such as depression, anxiety and chronic fatigue due to COVID-19 (Al-Jassas, Al-Hakeim et al. 2022). Indeed, TRYCATs are confirmed to be associated with various mental disorders, including depression, and anxiety (Maes, Mihaylova et al. 2007, Maes, Leonard et al. 2011), somatization and chronic fatigue syndrome (Maes and Rief 2012), cognitive impairments (Kanchanatawan, Hemrungrojn et al. 2018), and psychosis (Almulla, Vasupanrajit et al. 2021). Moreover, some TRYCATs, namely KYN, KA and 3HK are associated with musculoskeletal injuries due to their agonistic effects on the AhR (Duan and Lu 2019, Al Saedi, Sharma et al. 2020, Eisa, Reddy et al. 2020, Kondrikov, Elmansi et al. 2020). Thus, increased TRYCAT levels could exacerbate the neuro-immune and neuro-oxidative toxicity caused by increased oxidative stress and M1 and Th-1 activation resulting in comorbid affective disorders (Al-Jassas, Al-Hakeim et al. 2022). Therefore, it is safe to hypothesize that the accumulation of TRYCATs in SARS-CoV2 infected patients may play a role in the neuropsychiatric and cognitive syndromes of long or post-COVID syndrome (Taquet, Geddes et al. 2021).

Finally, it may be hypothesized that COVID-associated TRYCAT pathway activation may aggravate the disorders in the TRYCAT pathway in comorbid disorders (obesity, dementia, T2DM, hypertension and heart disease, stroke, chronic obstructive pulmonary disease (COPD) and chronic kidney disease), which increase risk to critical disease and death due to COVID-19 (Mayara Tiemi Enokida Mori 2021). Indeed, in all those comorbid diseases, the IDO enzyme is activated as indicated by an increased KYN/TRP ratio (Brouns, Verkerk et al. 2010, Gulaj, Pawlak et al. 2010, Mangge, Stelzer et al. 2014, Ormstad, Verkerk et al. 2014, Mallmann, Lima et al. 2018, Dschietzig, Kellner et al. 2019, Abedi, Vessal et al. 2021). By inference, when COVID-19 develops in people with those comorbid illnesses, an amplified TRYCATs response may occur, contributing to aggravated toxicity in addition to the consequences of inflammation and oxidative stress.

Another finding of our group meta-analysis revealed differences in the TRYCATs levels between COVID-19 patients and controls depending on whether plasma and serum was examined. For example, the results of KYN/TRP ratio in serum were highly significant with a large effect size (1.359), whereas in plasma no significant differences were found. Group analysis performed on the KA studies showed a significant difference in effect sizes between serum and plasma with serum KA yielding a positive medium effect size for COVID-19 (0.649), whereas in plasma a negative effect size was established. Similar results were detected in the associations between TRYCATs (e.g. KYN and KA) and schizophrenia with positive results in serum and often inverse results in plasma (Almulla, Vasupanrajit et al. 2021). The reader is referred to the latter paper for a discussion on the differences in TRYCAT measurements in serum versus plasma. Overall, it was concluded that TRYCATs measurements in serum are more adequate than assays performed in plasma (Almulla, Vasupanrajit et al. 2021).

Some limitations of the current systematic review and meta-analysis should be discussed. Not all studies clearly described the types of medications, the treatment protocol, the relevant comorbidities, and even the vaccination status of the patients. Moreover, non survivors following COVID-19 were sometimes lumped together with survivors. Due the small sample sizes and paucity of data on some TRYCATs, we were unable to estimate KMO and KYNU activity. Therefore, serum TRP and a more complete panel of serum TRYCATs should be determined in well-powered studies in the different stages of COVID-19 (i.e., mild, moderate, severe, critical, non-survival).

## Conclusions

Figure 1 summarizes the main findings of this study. The TRYCAT pathway is highly activated in COVID-19 and critical COVID-19 as indicated by increased IDO enzyme activity, which was assessed using the KYN/TRP ratio, and increased KYN but reduced TRP levels. KAT enzyme activity was not altered during COVID-19. These metabolites probably contribute to the pathophysiology of and severity and progression of COVID-19. ESF Table 6 shows the PRISMA checklist addressing all sections of our systematic review report.

## Data Availability

The utilized dataset (excel file) created to conduct this meta-analysis will be available from MM upon reasonable request and after the authors have completely exploited the dataset.

## Ethical approval and consent to participate

Not applicable.

## Consent for publication

Not applicable.

## Author’s contributions

This paper has been written with the contribution of all authors. AA and MM designed the work. Data collection was performed by AA and TS. AA and MM conducted the statistical analyses. The final draft was revised and approved by all authors.

## Acknowledgments

Not applicable.

## References

Abedi, S., M. Vessal, F. Asadian and M. A. Takhshid (2021). “Association of serum kynurenine/tryptophan ratio with poor glycemic control in patients with type2 diabetes.” Journal of Diabetes & Metabolic Disorders 20(2): 1521–1527.

Akbari, H., R. Tabrizi, K. B. Lankarani, H. Aria, S. Vakili, F. Asadian, S. Noroozi, P. Keshavarz and S. Faramarz (2020). “The role of cytokine profile and lymphocyte subsets in the severity of coronavirus disease 2019 (COVID-19): A systematic review and meta-analysis.” Life Sci 258: 118167.

Al-Jassas, H. K., H. K. Al-Hakeim and M. Maes (2022). “Intersections between pneumonia, lowered oxygen saturation percentage and immune activation mediate depression, anxiety, and chronic fatigue syndrome-like symptoms due to COVID-19: A nomothetic network approach.” Journal of Affective Disorders 297: 233–245.

Al Saedi, A., S. Sharma, M. A. Summers, K. Nurgali and G. Duque (2020). “The multiple faces of tryptophan in bone biology.” Exp Gerontol 129: 110778.

Almulla, A. F. and M. Maes (2022). “The tryptophan catabolite or kynurenine pathway’s role in major depression.” Preprints 2022010134.

Almulla, A. F., A. Vasupanrajit, C. Tunvirachaisakul, H. K. Al-Hakeim, M. Solmi, R. Verkerk and M. Maes (2021). “The tryptophan catabolite or kynurenine pathway in schizophrenia: meta-analysis reveals dissociations between central, serum and plasma compartments.” medRxiv: 2021.2012.2016.21267905.

Anderson, G., M. Maes and M. Berk (2013). “Schizophrenia is primed for an increased expression of depression through activation of immuno-inflammatory, oxidative and nitrosative stress, and tryptophan catabolite pathways.” Prog Neuropsychopharmacol Biol Psychiatry 42: 101–114.

Ansone, L., M. Briviba, I. Silamikelis, A. Terentjeva, I. Perkons, L. Birzniece, V. Rovite, B. Rozentale, L. Viksna, O. Kolesova, K. Klavins and J. Klovins (2021). “Amino Acid Metabolism is Significantly Altered at the Time of Admission in Hospital for Severe COVID-19 Patients: Findings from Longitudinal Targeted Metabolomics Analysis.” Microbiol Spectr 9(3): e0033821.

Blanco-Melo, D., B. E. Nilsson-Payant, W. C. Liu, S. Uhl, D. Hoagland, R. Møller, T. X. Jordan, K. Oishi, M. Panis, D. Sachs, T. T. Wang, R. E. Schwartz, J. K. Lim, R. A. Albrecht and B. R. tenOever (2020). “Imbalanced Host Response to SARS-CoV-2 Drives Development of COVID-19.” Cell 181(5): 1036–1045.e1039.

Blasco, H., C. Bessy, L. Plantier, A. Lefevre, E. Piver, L. Bernard, J. Marlet, K. Stefic, I. Benz-de Bretagne, P. Cannet, H. Lumbu, T. Morel, P. Boulard, C. R. Andres, P. Vourc’h, O. Herault, A. Guillon and P. Emond (2020). “The specific metabolome profiling of patients infected by SARS-COV-2 supports the key role of tryptophan-nicotinamide pathway and cytosine metabolism.” Sci Rep 10(1): 16824.

Bonaccorso, S., V. Marino, A. Puzella, M. Pasquini, M. Biondi, M. Artini, C. Almerighi, R. Verkerk, H. Meltzer and M. Maes (2002). “Increased depressive ratings in patients with hepatitis C receiving interferon-alpha-based immunotherapy are related to interferon-alpha-induced changes in the serotonergic system.” J Clin Psychopharmacol 22(1): 86–90.

Brosnahan, S. B., A. H. Jonkman, M. C. Kugler, J. S. Munger and D. A. Kaufman (2020). “COVID-19 and Respiratory System Disorders: Current Knowledge, Future Clinical and Translational Research Questions.” Arterioscler Thromb Vasc Biol 40(11): 2586–2597.

Brouns, R., R. Verkerk, T. Aerts, D. De Surgeloose, A. Wauters, S. Scharpé and P. P. De Deyn (2010). “The Role of Tryptophan Catabolism along the Kynurenine Pathway in Acute Ischemic Stroke.” Neurochemical Research 35(9): 1315–1322.

Cai, Y., D. J. Kim, T. Takahashi, D. I. Broadhurst, H. Yan, S. Ma, N. J. W. Rattray, A. Casanovas-Massana, B. Israelow, J. Klein, C. Lucas, T. Mao, A. J. Moore, M. C. Muenker, J. E. Oh, J. Silva, P. Wong, I. R. t. Yale, A. I. Ko, S. A. Khan, A. Iwasaki and C. H. Johnson (2021). “Kynurenic acid may underlie sex-specific immune responses to COVID-19.” Sci Signal 14(690).

Cohen, J. (2013). Statistical power analysis for the behavioral sciences, Academic press.

Coomes, E. A. and H. Haghbayan (2020). “Interleukin-6 in Covid-19: A systematic review and meta-analysis.” Rev Med Virol 30(6): 1–9.

D’Amora, P., I. Silva, M. A. Budib, R. Ayache, R. M. S. Silva, F. C. Silva, R. M. Appel, S. S. Junior, H. B. D. Pontes, A. C. Alvarenga, E. C. Arima, W. G. Martins, N. L. F. Silva, R. S. Diaz, M. B. Salzgeber, A. M. Palma, S. S. Evans and R. A. Nagourney (2021). “Towards risk stratification and prediction of disease severity and mortality in COVID-19: Next generation metabolomics for the measurement of host response to COVID-19 infection.” PLoS One 16(12): e0259909.

Doğan, S., T. Bal, M. Çabalak, N. Dikmen, H. Yaqoobi and O. Ozcan (2021). “Oxidative stress index can be a new marker related to disease severity in COVID-19.” Turkish Journal of Biochemistry 46(4): 349–357.

Dschietzig, T. B., K. H. Kellner, K. Sasse, F. Boschann, R. Klusener, J. Ruppert, F. P. Armbruster, D. Bankovic, A. Meinitzer, V. Mitrovic and C. Melzer (2019). “Plasma Kynurenine Predicts Severity and Complications of Heart Failure and Associates with Established Biochemical and Clinical Markers of Disease.” Kidney Blood Press Res 44(4): 765–776.

Duan, Z. and J. Lu (2019). “Involvement of Aryl Hydrocarbon Receptor in L-Kynurenine-Mediated Parathyroid Hormone-Related Peptide Expression.” Horm Cancer 10(2-3): 89–96.

Dykens, J. A., S. G. Sullivan and A. Stern (1987). “Oxidative reactivity of the tryptophan metabolites 3-hydroxyanthranilate, cinnabarinate, quinolinate and picolinate.” Biochem Pharmacol 36(2): 211–217.

Eisa, N. H., S. V. Reddy, A. M. Elmansi, G. Kondrikova, D. Kondrikov, X. M. Shi, C. M. Novince, M. W. Hamrick, M. E. McGee-Lawrence, C. M. Isales, S. Fulzele and W. D. Hill (2020). “Kynurenine Promotes RANKL-Induced Osteoclastogenesis In Vitro by Activating the Aryl Hydrocarbon Receptor Pathway.” Int J Mol Sci 21(21).

Fallarino, F., U. Grohmann, C. Vacca, R. Bianchi, C. Orabona, A. Spreca, M. C. Fioretti and P. Puccetti (2002). “T cell apoptosis by tryptophan catabolism.” Cell Death Differ 9(10): 1069–1077.

Fallarino, F., U. Grohmann, S. You, B. C. McGrath, D. R. Cavener, C. Vacca, C. Orabona, R. Bianchi, M. L. Belladonna, C. Volpi, P. Santamaria, M. C. Fioretti and P. Puccetti (2006). “The combined effects of tryptophan starvation and tryptophan catabolites down-regulate T cell receptor zeta-chain and induce a regulatory phenotype in naive T cells.” J Immunol 176(11): 6752–6761.

Fraser, D. D., M. Slessarev, C. M. Martin, M. Daley, M. A. Patel, M. R. Miller, E. K. Patterson, D. B. O’Gorman, S. E. Gill, D. S. Wishart, R. Mandal and G. Cepinskas (2020). “Metabolomics Profiling of Critically Ill Coronavirus Disease 2019 Patients: Identification of Diagnostic and Prognostic Biomarkers.” Crit Care Explor 2(10): e0272.

Gadotti, A. C., M. de Castro Deus, J. P. Telles, R. Wind, M. Goes, R. Garcia Charello Ossoski, A. M. de Padua, L. de Noronha, A. Moreno-Amaral, C. P. Baena and F. F. Tuon (2020). “IFN-γ is an independent risk factor associated with mortality in patients with moderate and severe COVID-19 infection.” Virus research 289: 198171–198171.

Giovannoni, F., Z. Li, C. C. Garcia and F. J. Quintana (2020). “A potential role for AHR in SARS-CoV-2 pathology.” Res Sq.

Goda, K., Y. Hamane, R. Kishimoto and Y. Ogishi (1999). “Radical scavenging properties of tryptophan metabolites. Estimation of their radical reactivity.” Adv Exp Med Biol 467: 397–402.

Goldstein, L. E., M. C. Leopold, X. Huang, C. S. Atwood, A. J. Saunders, M. Hartshorn, J. T. Lim, K. Y. Faget, J. A. Muffat, R. C. Scarpa, L. T. Chylack, Jr., E. F. Bowden, R. E. Tanzi and A. I. Bush (2000). “3-Hydroxykynurenine and 3-hydroxyanthranilic acid generate hydrogen peroxide and promote alpha-crystallin cross-linking by metal ion reduction.” Biochemistry 39(24): 7266–7275.

Guidetti, P. and R. Schwarcz (1999). “3-Hydroxykynurenine potentiates quinolinate but not NMDA toxicity in the rat striatum.” Eur J Neurosci 11(11): 3857–3863.

Guillemin, G. J., K. M. Cullen, C. K. Lim, G. A. Smythe, B. Garner, V. Kapoor, O. Takikawa and B. J. Brew (2007). “Characterization of the kynurenine pathway in human neurons.” J Neurosci 27(47): 12884–12892.

Gulaj, E., K. Pawlak, B. Bien and D. Pawlak (2010). “Kynurenine and its metabolites in Alzheimer’s disease patients.” Advances in Medical Sciences 55(2): 204–211.

Herrera-Van Oostdam, A. S., J. E. Castaneda-Delgado, J. J. Oropeza-Valdez, J. C. Borrego, J. Monarrez-Espino, J. Zheng, R. Mandal, L. Zhang, E. Soto-Guzman, J. C. Fernandez-Ruiz, F. Ochoa-Gonzalez, F. M. Trejo Medinilla, J. A. Lopez, D. S. Wishart, J. A. Enciso-Moreno and Y. Lopez-Hernandez (2021). “Immunometabolic signatures predict risk of progression to sepsis in COVID-19.” PLoS One 16(8): e0256784.

Higgins JPT, T. J., Chandler J, Cumpston M, Li T, Page MJ, Welch VA (2019). Cochrane Handbook for Systematic Reviews of Interventions. Chichester (UK), John Wiley & Sons.

Hojyo, S., M. Uchida, K. Tanaka, R. Hasebe, Y. Tanaka, M. Murakami and T. Hirano (2020). “How COVID-19 induces cytokine storm with high mortality.” Inflamm Regen 40:37.

Kanchanatawan, B., S. Hemrungrojn, S. Thika, S. Sirivichayakul, K. Ruxrungtham, A. F. Carvalho, M. Geffard, G. Anderson and M. Maes (2018). “Changes in Tryptophan Catabolite (TRYCAT) Pathway Patterning Are Associated with Mild Impairments in Declarative Memory in Schizophrenia and Deficits in Semantic and Episodic Memory Coupled with Increased False-Memory Creation in Deficit Schizophrenia.” Mol Neurobiol 55(6): 5184–5201.

Kanchanatawan, B., S. Sirivichayakul, K. Ruxrungtham, A. F. Carvalho, M. Geffard, H. Ormstad, G. Anderson and M. Maes (2018). “Deficit, but Not Nondeficit, Schizophrenia Is Characterized by Mucosa-Associated Activation of the Tryptophan Catabolite (TRYCAT) Pathway with Highly Specific Increases in IgA Responses Directed to Picolinic, Xanthurenic, and Quinolinic Acid.” Mol Neurobiol 55(2): 1524–1536.

Kimhofer, T., S. Lodge, L. Whiley, N. Gray, R. L. Loo, N. G. Lawler, P. Nitschke, S. H. Bong, D. L. Morrison, S. Begum, T. Richards, B. B. Yeap, C. Smith, K. G. C. Smith, E. Holmes and J. K. Nicholson (2020). “Integrative Modeling of Quantitative Plasma Lipoprotein, Metabolic, and Amino Acid Data Reveals a Multiorgan Pathological Signature of SARS-CoV-2 Infection.” J Proteome Res 19(11): 4442–4454.

Kondrikov, D., A. Elmansi, R. T. Bragg, T. Mobley, T. Barrett, N. Eisa, G. Kondrikova, P. Schoeinlein, A. Aguilar-Perez, X. M. Shi, S. Fulzele, M. M. Lawrence, M. Hamrick, C. Isales and W. Hill (2020). “Kynurenine inhibits autophagy and promotes senescence in aged bone marrow mesenchymal stem cells through the aryl hydrocarbon receptor pathway.” Exp Gerontol 130: 110805.

Laforge, M., C. Elbim, C. Frere, M. Hemadi, C. Massaad, P. Nuss, J. J. Benoliel and C. Becker (2020). “Tissue damage from neutrophil-induced oxidative stress in COVID-19.” Nat Rev Immunol 20(9): 515–516.

Lawler, N. G., N. Gray, T. Kimhofer, B. Boughton, M. Gay, R. Yang, A. C. Morillon, S. T. Chin, M. Ryan, S. Begum, S. H. Bong, J. D. Coudert, D. Edgar, E. Raby, S. Pettersson, T. Richards, E. Holmes, L. Whiley and J. K. Nicholson (2021). “Systemic Perturbations in Amine and Kynurenine Metabolism Associated with Acute SARS-CoV-2 Infection and Inflammatory Cytokine Responses.” J Proteome Res 20(5): 2796–2811.

Lee, G. K., H. J. Park, M. Macleod, P. Chandler, D. H. Munn and A. L. Mellor (2002). “Tryptophan deprivation sensitizes activated T cells to apoptosis prior to cell division.” Immunology 107(4): 452–460.

Lionetto, L., M. Ulivieri, M. Capi, D. De Bernardini, F. Fazio, A. Petrucca, L. M. Pomes, O. De Luca, G. Gentile, B. Casolla, M. Curto, G. Salerno, S. Schillizzi, M. S. Torre, I. Santino, M. Rocco, P. Marchetti, A. Aceti, A. Ricci, R. Bonfini, F. Nicoletti, M. Simmaco and M. Borro (2021). “Increased kynurenine-to-tryptophan ratio in the serum of patients infected with SARS-CoV2: An observational cohort study.” Biochim Biophys Acta Mol Basis Dis 1867(3): 166042.

Lugo-Huitrón, R., T. Blanco-Ayala, P. Ugalde-Muñiz, P. Carrillo-Mora, J. Pedraza-Chaverrí, D. Silva-Adaya, P. D. Maldonado, I. Torres, E. Pinzón, E. Ortiz-Islas, T. López, E. García, B. Pineda, M. Torres-Ramos, A. Santamaría and V. P. La Cruz (2011). “On the antioxidant properties of kynurenic acid: free radical scavenging activity and inhibition of oxidative stress.” Neurotoxicol Teratol 33(5): 538–547.

Lugo-Huitron, R., P. Ugalde Muniz, B. Pineda, J. Pedraza-Chaverri, C. Rios and V. Perez-de la Cruz (2013). “Quinolinic acid: an endogenous neurotoxin with multiple targets.” Oxid Med Cell Longev 2013: 104024.

Maes, M. and G. Anderson (2021). “False Dogmas in Schizophrenia Research: Toward the Reification of Pathway Phenotypes and Pathway Classes.” 12(963).

Maes, M., B. E. Leonard, A. M. Myint, M. Kubera and R. Verkerk (2011). “The new ’5-HT’ hypothesis of depression: cell-mediated immune activation induces indoleamine 2,3-dioxygenase, which leads to lower plasma tryptophan and an increased synthesis of detrimental tryptophan catabolites (TRYCATs), both of which contribute to the onset of depression.” Prog Neuropsychopharmacol Biol Psychiatry 35(3): 702–721.

Maes, M., B. E. Leonard, A. M. Myint, M. Kubera and R. Verkerk (2011). “The new ‘5-HT’ hypothesis of depression: Cell-mediated immune activation induces indoleamine 2,3-dioxygenase, which leads to lower plasma tryptophan and an increased synthesis of detrimental tryptophan catabolites (TRYCATs), both of which contribute to the onset of depression.” Progress in Neuro-Psychopharmacology and Biological Psychiatry 35(3): 702–721.

Maes, M., I. Mihaylova, M. D. Ruyter, M. Kubera and E. Bosmans (2007). “The immune effects of TRYCATs (tryptophan catabolites along the IDO pathway): relevance for depression - and other conditions characterized by tryptophan depletion induced by inflammation.” Neuro Endocrinol Lett 28(6): 826–831.

Maes, M. and W. Rief (2012). “Diagnostic classifications in depression and somatization should include biomarkers, such as disorders in the tryptophan catabolite (TRYCAT) pathway.” Psychiatry Res 196(2-3): 243–249.

Maes, M., W. L. D. Tedesco Junior, M. A. B. Lozovoy, M. T. E. Mori, T. Danelli, E. R. D. Almeida, A. M. Tejo, Z. N. Tano, E. M. V. Reiche and A. N. C. Simao (2022). “In COVID-19, NLRP3 inflammasome genetic variants are associated with critical disease and these effects are partly mediated by the sickness symptom complex: a nomothetic network approach.” Mol Psychiatry.

Mallmann, N. H., E. S. Lima and P. Lalwani (2018). “Dysregulation of Tryptophan Catabolism in Metabolic Syndrome.” Metab Syndr Relat Disord 16(3): 135–142.

Mandi, Y. and L. Vecsei (2012). “The kynurenine system and immunoregulation.” J Neural Transm (Vienna) 119(2): 197–209.

Mangge, H., M. Herrmann, A. Meinitzer, S. Pailer, P. Curcic, Z. Sloup, M. Holter and F. Pruller (2021). “Increased Kynurenine Indicates a Fatal Course of COVID-19.” Antioxidants (Basel) 10(12).

Mangge, H., I. Stelzer, E. Z. Reininghaus, D. Weghuber, T. T. Postolache and D. Fuchs (2014). “Disturbed tryptophan metabolism in cardiovascular disease.” Current medicinal chemistry 21(17): 1931–1937.

Marin-Corral, J., J. Rodriguez-Morato, A. Gomez-Gomez, S. Pascual-Guardia, R. Munoz-Bermudez, A. Salazar-Degracia, P. Perez-Teran, M. I. Restrepo, O. Khymenets, N. Haro, J. R. Masclans and O. J. Pozo (2021). “Metabolic Signatures Associated with Severity in Hospitalized COVID-19 Patients.” Int J Mol Sci 22(9).

Mayara Tiemi Enokida Mori, A. N. C. S., Tiago Danelli, Sayonara Rangel Oliveira, Pedro Luis Candido de Souza Cassela, Guilherme Lerner Trigo, Kauê Cardoso, Alexandre Mestre Tejo, Zuleica Naomi Tanno, Elaine Regina Delicato de Almeida, Edna Maria Vissoci Reiche, Michael Maes, Marcell Alysson Batisti Lozovoy (2021). “Protective effects of IL18-105G>A and IL18-137C>G genetic variants on severity of COVID-19.” <u>Preprint</u>.

Mellor, A. L. and D. H. Munn (1999). “Tryptophan catabolism and T-cell tolerance: immunosuppression by starvation?” Immunology Today 20(10): 469–473.

Michaelis, S., S. Zelzer, W. J. Schnedl, A. Baranyi, A. Meinitzer and D. Enko (2021). “Assessment of tryptophan and kynurenine as prognostic markers in patients with SARS-CoV-2.” Clin Chim Acta 525: 29–33.

Mohiuddin, M. and K. Kasahara (2021). “The emerging role of oxidative stress in complications of COVID-19 and potential therapeutic approach to diminish oxidative stress.” Respir Med 187: 106605.

Morris, G., A. F. Carvalho, G. Anderson, P. Galecki and M. Maes (2016). “The Many Neuroprogressive Actions of Tryptophan Catabolites (TRYCATs) that may be Associated with the Pathophysiology of Neuro-Immune Disorders.” Curr Pharm Des 22(8): 963–977.

Muhoberac, B. B. (2020). “What Can Cellular Redox, Iron, and Reactive Oxygen Species Suggest About the Mechanisms and Potential Therapy of COVID-19?” Front Cell Infect Microbiol 10: 569709.

Mulchandani, R., T. Lyngdoh and A. K. Kakkar (2021). “Deciphering the COVID-19 cytokine storm: Systematic review and meta-analysis.” Eur J Clin Invest 51(1): e13429.

Okuda, S., N. Nishiyama, H. Saito and H. Katsuki (1998). “3-Hydroxykynurenine, an endogenous oxidative stress generator, causes neuronal cell death with apoptotic features and region selectivity.” J Neurochem 70(1): 299–307.

Ombrello, M. J. and G. S. Schulert (2021). “COVID-19 and cytokine storm syndrome: are there lessons from macrophage activation syndrome?” Transl Res 232: 1–12.

Ormstad, H., R. Verkerk, K. F. Amthor and L. Sandvik (2014). “Activation of the kynurenine pathway in the acute phase of stroke and its role in fatigue and depression following stroke.” J Mol Neurosci 54(2): 181–187.

Page, M. J., J. E. McKenzie, P. M. Bossuyt, I. Boutron, T. C. Hoffmann, C. D. Mulrow, L. Shamseer, J. M. Tetzlaff, E. A. Akl, S. E. Brennan, R. Chou, J. Glanville, J. M. Grimshaw, A. Hrobjartsson, M. M. Lalu, T. Li, E. W. Loder, E. Mayo-Wilson, S. McDonald, L. A. McGuinness, L. A. Stewart, J. Thomas, A. C. Tricco, V. A. Welch, P. Whiting and D. Moher (2021). “The PRISMA 2020 statement: an updated guideline for reporting systematic reviews.” BMJ 372: n71.

Page, M. J., J. E. McKenzie, P. M. Bossuyt, I. Boutron, T. C. Hoffmann, C. D. Mulrow, L. Shamseer, J. M. Tetzlaff, E. A. Akl, S. E. Brennan, R. Chou, J. Glanville, J. M. Grimshaw, A. Hróbjartsson, M. M. Lalu, T. Li, E. W. Loder, E. Mayo-Wilson, S. McDonald, L. A. McGuinness, L. A. Stewart, J. Thomas, A. C. Tricco, V. A. Welch, P. Whiting and D. Moher (2021). “The PRISMA 2020 statement: An updated guideline for reporting systematic reviews.” PLoS medicine 18(3): e1003583–e1003583.

Pakorn Sagulkoo, K. P., Apichat Suratanee, Andrea Name Colado Simão, Edna Maria Vissoci Reiche, Michael Maes (2022). “Immunopathogenesis and immunogenetic variants in COVID-19.” Preprint.

Pallotta, M. T., F. Fallarino, D. Matino, A. Macchiarulo and C. Orabona (2014). “AhR-Mediated, Non-Genomic Modulation of IDO1 Function.” Front Immunol 5: 497.

Pérez-González, A., J. R. Alvarez-Idaboy and A. Galano (2015). “Free-radical scavenging by tryptophan and its metabolites through electron transfer based processes.” J Mol Model 21(8): 213.

Ragab, D., H. Salah Eldin, M. Taeimah, R. Khattab and R. Salem (2020). “The COVID-19 Cytokine Storm; What We Know So Far.” Front Immunol 11: 1446.

Rahman, A., M. S. Rao and K. M. Khan (2018). “Intraventricular infusion of quinolinic acid impairs spatial learning and memory in young rats: a novel mechanism of lead-induced neurotoxicity.” J Neuroinflammation 15(1): 263.

Reyes Ocampo, J., R. Lugo Huitrón, D. González-Esquivel, P. Ugalde-Muñiz, A. Jiménez-Anguiano, B. Pineda, J. Pedraza-Chaverri, C. Ríos and V. Pérez de la Cruz (2014). “Kynurenines with Neuroactive and Redox Properties: Relevance to Aging and Brain Diseases.” Oxidative Medicine and Cellular Longevity 2014: 646909.

Robertson, J., J. M. Gostner, S. Nilsson, L. M. Andersson, D. Fuchs and M. Gisslen (2020). “Serum neopterin levels in relation to mild and severe COVID-19.” BMC Infect Dis 20(1):942.

Santamaría, A., S. Galván-Arzate, V. Lisý, S. F. Ali, H. M. Duhart, L. Osorio-Rico, C. Ríos and F. St’astný (2001). “Quinolinic acid induces oxidative stress in rat brain synaptosomes.” Neuroreport 12(4): 871–874.

Schmidt, S. K., A. Muller, K. Heseler, C. Woite, K. Spekker, C. R. MacKenzie and W. Daubener (2009). “Antimicrobial and immunoregulatory properties of human tryptophan 2,3-dioxygenase.” Eur J Immunol 39(10): 2755–2764.

Shen, B., X. Yi, Y. Sun, X. Bi, J. Du, C. Zhang, S. Quan, F. Zhang, R. Sun, L. Qian, W. Ge, W. Liu, S. Liang, H. Chen, Y. Zhang, J. Li, J. Xu, Z. He, B. Chen, J. Wang, H. Yan, Y. Zheng, D. Wang, J. Zhu, Z. Kong, Z. Kang, X. Liang, X. Ding, G. Ruan, N. Xiang, X. Cai, H. Gao, L. Li, S. Li, Q. Xiao, T. Lu, Y. Zhu, H. Liu, H. Chen and T. Guo (2020). “Proteomic and Metabolomic Characterization of COVID-19 Patient Sera.” Cell 182(1):59–72 e15.

Smith, A. J., R. A. Smith and T. W. Stone (2009). “5-Hydroxyanthranilic acid, a tryptophan metabolite, generates oxidative stress and neuronal death via p38 activation in cultured cerebellar granule neurones.” Neurotox Res 15(4): 303–310.

Smith, R. S. and M. Maes (1995). “The macrophage-T-lymphocyte theory of schizophrenia: additional evidence.” Med Hypotheses 45(2): 135–141.

Taquet, M., J. R. Geddes, M. Husain, S. Luciano and P. J. Harrison (2021). “6-month neurological and psychiatric outcomes in 236 379 survivors of COVID-19: a retrospective cohort study using electronic health records.” The Lancet Psychiatry 8(5): 416–427.

Thomas, T., D. Stefanoni, J. A. Reisz, T. Nemkov, L. Bertolone, R. O. Francis, K. E. Hudson, J. C. Zimring, K. C. Hansen, E. A. Hod, S. L. Spitalnik and A. D’Alessandro (2020). “COVID-19 infection alters kynurenine and fatty acid metabolism, correlating with IL-6 levels and renal status.” JCI Insight 5(14).

Turski, W. A., A. Wnorowski, G. N. Turski, C. A. Turski and L. Turski (2020). “AhR and IDO1 in pathogenesis of Covid-19 and the “Systemic AhR Activation Syndrome:” a translational review and therapeutic perspectives.” Restor Neurol Neurosci 38(4): 343–354.

Vasupanrajit, A., K. Jirakran, C. Tunvirachaisakul and M. Maes (2021). “Suicide attempts are associated with activated immune-inflammatory, nitro-oxidative, and neurotoxic pathways: A systematic review and meta-analysis.” Journal of Affective Disorders 295:80–92.

Vasupanrajit, A., K. Jirakran, C. Tunvirachaisakul, M. Solmi and M. Maes (2021). “Inflammation and nitro-oxidative stress in current suicidal attempts and current suicidal ideation: a systematic review and meta-analysis.” medRxiv: 2021.2009.2009.21263363.

Vora, S. M., J. Lieberman and H. Wu (2021). “Inflammasome activation at the crux of severe COVID-19.” Nat Rev Immunol 21(11): 694–703.

Vyavahare, S., S. Kumar, N. Cantu, R. Kolhe, W. B. Bollag, M. E. McGee-Lawrence, W. D. Hill, M. W. Hamrick, C. M. Isales and S. Fulzele (2021). “Tryptophan-Kynurenine Pathway in COVID-19-Dependent Musculoskeletal Pathology: A Minireview.” Mediators Inflamm 2021: 2911578.

Wan, X., W. Wang, J. Liu and T. Tong (2014). “Estimating the sample mean and standard deviation from the sample size, median, range and/or interquartile range.” BMC Medical Research Methodology 14(1): 135.

Wirthgen, E., A. Hoeflich, A. Rebl and J. Günther (2018). “Kynurenic Acid: The Janus-Faced Role of an Immunomodulatory Tryptophan Metabolite and Its Link to Pathological Conditions.” Frontiers in Immunology 8.

Xiao, N., M. Nie, H. Pang, B. Wang, J. Hu, X. Meng, K. Li, X. Ran, Q. Long, H. Deng, N. Chen, S. Li, N. Tang, A. Huang and Z. Hu (2021). “Integrated cytokine and metabolite analysis reveals immunometabolic reprogramming in COVID-19 patients with therapeutic implications.” Nat Commun 12(1): 1618.

Xu, H., G. X. Zhang, B. Ciric and A. Rostami (2008). “IDO: a double-edged sword for T(H)1/T(H)2 regulation.” Immunol Lett 121(1): 1–6.

Yan, Y., G. X. Zhang, B. Gran, F. Fallarino, S. Yu, H. Li, M. L. Cullimore, A. Rostami and H. Xu (2010). “IDO upregulates regulatory T cells via tryptophan catabolite and suppresses encephalitogenic T cell responses in experimental autoimmune encephalomyelitis.” J Immunol 185(10): 5953–5961.

Yang, L., X. Xie, Z. Tu, J. Fu, D. Xu and Y. Zhou (2021). “The signal pathways and treatment of cytokine storm in COVID-19.” Signal Transduct Target Ther 6(1): 255.

Yin, K., E. Gribbin and H. Wang (2005). “Interferon-gamma inhibition attenuates lethality after cecal ligation and puncture in rats: implication of high mobility group box-1.” Shock 24(4): 396–401.

