## supplementary file for "The tryptophan catabolite or kynurenine pathway in COVID-19 and critical COVID-19: a systematic review and meta-analysis"

SHORTTITLE: Kynurenine pathway in COVID-19

Abbas F. Almulla, Ph.D.^a,b^ , Thitiporn Supasitthumrong, M.D., Ph.D.^a^, Chavit Tunvirachaisakul, M.D., Ph.D.^a^, Ali Abbas Abo Algon, MS.c ^c^, Hussein K. Al-Hakeim, Ph.D.^d^, Michael Maes, M.D., Ph.D.^a,e,f^

^a^ Department of Psychiatry, Faculty of Medicine, Chulalongkorn University, Bangkok, Thailand.

^b^ Medical Laboratory Technology Department, College of Medical Technology, The Islamic University, Najaf, Iraq.

^c^ Iraqi Education Ministry- Najaf- Iraq.

^d^ Department of Chemistry, College of Science, University of Kufa, Kufa, Iraq.

^e^ Department of Psychiatry, Medical University of Plovdiv, Plovdiv, Bulgaria.

^f^ Department of Psychiatry, IMPACT Strategic Research Centre, Deakin University, Geelong, Victoria, Australia.

**ESF, Table 1:** Search sentences and terms used in each database

| **Database Name** | **Search Sentence** | **No. of Articles** |
| --- | --- | --- |
| **PubMed/Medline** | (((((((((((((COVID-10 and TRYCATs)* OR (COVID-19 and kynurenine)) OR (COVID-19 and kynurenic acid)) OR (COVID-19 and quinolinic acid)) OR (COVID-19 and Picolinic acid)) OR (COVID-19 and 3-hydroxyanthranilic acid)) OR (COVID-19 and xanthurenic acid)) OR (COVID-19 and 3-hydroxykynurenine)) OR (COVID-19 and Anthranilic acid)) OR (COVID-19 and formyl kynurenine)) OR (COVID-19 and l-tryptophan)) OR (COVID-19 and tryptophan catabolites)) OR (TRYCATs and COVID-19)) OR (TRYCATs and Coronavirus) | **106** |
|  | (((((((((((TRYCATs and COVID-19) OR (TRYCATs and SARS-CoV-2)) OR (Tryptophan Catabolites and COVID-19)) OR (L-TRP and COVID-19)) OR (L-KYN and COVID-19)) OR (KYNA and COVID-19)) OR (ANA and COVID-19)) OR (3-HK and COVID-19)) OR (XA and COVID-19)) OR (3-HANA and COVID-19)) OR (QA and COVID-19)) OR (PA and COVID-19) | **106** |
|  | ((((((((((SARS-CoV-2 and TRYCATs)* OR (severe acute respiratory syndrome coronavirus 2 and Tryptophan)) OR (COVID-19 and tryptophan cata)) OR (severe acute respiratory syndrome coronavirus 2 and TRYCATs)) OR (SARS-CoV-2 and TRP)) OR (SARS-CoV-2 and KYN)) OR (SARS-CoV-2 and kynurenine)) OR (SARS-CoV-2 and Kynurenine pathway)) OR (COVID-19 and kynurenine pathway)) OR (COVID-19 and IDO)) OR (COVID-19 and TDO) | **110** |
| **Google Scholar** | (((((COVID-19* and IDO activation) OR (TDO activation)) OR (Kynurenine pathway)) OR (KMO activation)) OR (Tryptophan degradation)) AND (Decreased Tryptophan) | **395** |
| **Web of Science** | (((((((ALL=(COVID-19, TRYCATs)) OR ALL=(COVID-19, Tryptophan catabolites )) OR ALL=(COVID-19, kynurenine pathway )) OR ALL=(COVID-19, IDO and TDO activity )) OR ALL=(COVID-19, xanthurenic acid, picolinic acid))) OR ALL=(COVID-19, kynurenic acid and kynurenine )) OR ALL=(COVID-19, anthranilic acid and 3-hydroxyanthranlinic acid ) | **23** |

**ESF, Table 2.** Immune cofounder’s scale (ICS) (adapted from Andrés-Rodríguez, et al., 2019)

| **Methodological quality of the study** | |
| --- | --- |
| **1** | Study sample ≥ 128 participants including patients and controls (1= Yes, 0 = No) |
| **2** | Did the study control the results for potential confounders (e.g., age, BMI, gender, race)? (1= Yes, 0 = No) |
| **3** | Were participants with COVID-19 patients and controls age- and-gender-matched or was there a statistical control? (1= Yes, 0 = No) |
| **4** | Was the time of sample collection specified (e.g., morning vs. evening)? (1= Yes, 0 = No) |
| **5** | Were participants with COVID-19 free of immunomodulatory drugs including anti-cytokines, glucocorticoids, immunoglobulins, and immunosuppressants, or was there a medication washout period or was drug intake statistically controlled for? (1= Yes, 0 = No) |
| **6** | Were participants with COVID-19 free of medications or were the data statistically controlled for? (1= Yes, 0 = No) |
| **7** | Reporting either the manufacturer of the test or detection limit and coefficients of variation (1= Yes, 0 = No) |
| **8** | Reporting how data under detection limit were handled (1 = Yes, 0 = No) |
| **9** | Reporting % of the sample under detection limit (1=Yes, 0= No) |
| **10** | Reporting blood fraction (serum, plasma, culture supernatant or whole blood) (1= Yes, 0 = No) |
| **Total quality score (10 points)** | |
| **Biomarker confounders red points**  *The red points should not be given if the item is statistically controlled for* | |
| **1** | 3 red points for comorbid illnesses such as autoimmune disorders & other immune disorders including rheumatoid arthritis, psoriasis, inflammatory bowel disease, chronic obstructive pulmonary disease, multiple sclerosis |
| **2** | 3 red points for use of recreational drugs such as methamphetamine or opioids |
| **3** | 3 red point for use of omega-3 and antioxidant supplements |
| **4** | 3 red point for more common systemic immune disorders including diabetes type 1/2, essential hypertension, metabolic syndrome |
| **5** | 2 red points when groups were not matched for age |
| **6** | 2 red points when groups were not matched for sex |
| **7** | 2 red points for medication use as for example immunomodulators |
| **8** | 2 red point when data were not controlled for smoking |
| **9** | 2 red point when data were not controlled for body mass index |
| **10** | 1 red point for not fasting (8 hours before blood extraction) |
| **11** | 1 red point for use of oral contraceptives or NSAIDs |
| **12** | 1 red points when data were not controlled for ethnicity in countries such as US, Brazil |
| **13** | 1 red points when data were not controlled for diurnal variation (8-10 a.m. versus all other time points) |
|  | **Total red point score (26 points)** |

**ESF. Table 3.** Studies excluded from the meta-analysis but included in the systematic review.

| **Authors, year** | **Reason why excluded from the meta-analysis** |
| --- | --- |
| Cai, Kim et al. 2020 | No Mean(SD) or any way to estimate them |
| Kaur, Ji et al. 2021 | No Mean(SD) or any way to estimate them |
| Lopez-Hernandez, Monarrez-Espino et al. 2021 | No Mean(SD) or any way to estimate them |
| Guo, Schurink et al. 2021 | No Mean(SD) or any way to estimate them |
| Danlos, Grajeda-Iglesias et al. 2021 | No Mean(SD) or any way to estimate them |

| **ESF, Table 4.** Characterstics of the studies included in the systematic reviews and meta-analysis | | | | | | | | | | | | | | | | |
| --- | --- | --- | --- | --- | --- | --- | --- | --- | --- | --- | --- | --- | --- | --- | --- | --- |
| **NO** | **Authors, years** | **Setting** | **Type of case** | **Type of Control** | **Sample Size** | | | **Age** | | **Assessed**  **Biomarkers** | **Specimen** | **Method** | **Severity** | **Quality score** | **Red point score** | **Findnigs** |
|  |  |  |  |  | **Cases**  **M/F** | **Control**  **M/F** | **Total**  **M/F** | **Case-Mean (SD)** | **Control- Mean(SD)** |  |  |  |  |  |  |  |
| **Studies were included in systematic review and meta-analysis** | | | | | | | | | | | | | | | | |
| 1 | Ansone, Briviba et al. 2021 | Latvia | +Covid-19 | HC | 33  16/17 | 39  16/23 | 72  32/40 | 58.5(16.2) | 53.4(14.7) | KYN, TRP | Serum | LC-MS | 1 | 6 | 13 | High KYN, Low TRP, High 3HK |
| 2 | Blasco, Bessy et al. 2020 | France | +Covid-19 | -Covid-19 | 55  27/28 | 45  23/22 | 100  50/50 | 77,5(16) | 75,9(17,5) | KYN, KA, TRP | Plasma | LC-HRMS | 1 | 3 | 6 | High KYN, Low TRP and KA |
| 3 | D'Amora, Silva et al. 2021 | Brazil | +Covid-19 | HC | 82  48/34 | 31  10/21 | 113  58/55 | 53.4(11.0) | 39,2(10,2) | KYN, TRP | Plasma | LC-MS | 1 | 3 | 10 | High KYN, Low TRP |
| 4 | Fraser, Slessarev et al. 2020 | Canada | +Covid-19 | HC | 10  7/3 | 10  7/3 | 20  10/10 | 60,9(10,4) | 57,7(8,6) | KYN | Plasma | LC-MS | 2 | 4 | 12 | High KYN |
| 5 | Herrera-Van Oostdam et al. 2021 | Mexico | +Covid-19 | -Covid-19 | 121  68/53 | 37  16/21 | 158  84/74 | 55.8(11.8) | 44,5(12,3) | KYN, TRP | Plasma | LC-MS/MS | 1 | 7 | 11 | High KYN, Low TRP |
| 6 | Kimhofer, Lodge et al. 2020 | Australia | +Covid-19 | HC | 17  11/6 | 25  17/8 | 42  28/14 | 69,4(10,5) | 48,8(15,3) | KYN, TRP | Plasma | LC-MS | 1 | 3 | 12 | Low KYN/TRP |
| 7 | Lawler, Gray et al. 2021 | Australia | +Covid-19 | HC | 10  7/3 | 16  10/6 | 26  17/9 | 68,4(12,6) | 50,3(16,7) | KYN,KA,3HK,QA,XA,PA | Plasma | UHPLC-MS | NA | 3 | 13 | High KYN, Low TRP, High KA |
| 8 | Lionetto, Ulivieri et al. 2021 | Italy | +Covid-19 | HC | 89  43/46 | 239  87/152 | 328  130/198 | 66(16,5) | 45,6(11,1) | KYN, TRP | Serum | LC–MS/MS | 1 | 4 | 14 | High KYN, Low TRP |
| 9 | Thomas, Stefanoni et al. 2020 | USA | +Covid-19 | -Covid-19 | 33  25/8 | 16  6/10 | 49  31/18 | 56,5(18,1) | 37,8(11,6) | KYN, KA, TRP | Serum | UHPLC-MS | 1 | 3 | 17 | High KYN, Low TRP, High KA |
| 10 | Xiao, Nie et al. 2021 | China | +Covid-19 | HC | 44  NA | 17  NA | 61  NA | NA | NA | KYN,KA, TRP | Serum | LC-MS | 1 | 3 | 12 | High KYN, Low TRP, High KA |
| 11 | Marin-Corral, Rodriguez-Morato et al. 2021 | Spain | Severe-Critical +Covid-19 | Moderate +Covid-19 | 36  17/19 | 13  7/6 | 49  24/25 | 59(10)  50(14) | 51(18) | TRP, KYN, 3HK | Plasma | LC–MS/MS | 2 | 3 | 10 | High KYN and 3Hk, Low TRP |
| 12 | Mangge, Herrmann et al. 2021 | Austria | Died +Covid-19 | Recoveried  +Covid-19 | 31  17/14 | 117  62/55 | 148  79/69 | 78.8(11.1) | 56.8(14.6) | KYN | Plasma | LC-UV | 2 | 4 | 13 | High KYN |
| 13 | Michaelis, Zelzer et al. 2021 | Austria | Died +Covid-19 | Survived +Covid-19 | 53  25/28 | 199  118/81 | 252 | 82(10.6) | 73.2(13.4) | TRP, KYN | Plasma | HPLC | 2 | 4 | 13 | High KYN, Low TRP |
| 14 | Robertson, Gostner et al. 2020 | Sweden | Severe +Covid-19 | Mild +Covid-19 | 19  17/2 | 15  6/9 | 34  23/11 | 61.3(11.4) | 51.3(13.7) | TRP, KYN | Serum | HPLC | 2 | 3 | 13 | High KYN, Low TRP |
| **Studies were included only in systematic review** | | | | | | | | | | | | | | | | |
| 1 | Cai, Kim et al. 2020 | USA | +Covid-19 | HC | 39 17/22 | 20 10/10 | 59  27/32 | NA | NA | Serum | KA, KYN | (HILIC)–MS and (RPLC)–MS | 1 | 3 | 14 | High KA/KYN ratio |
| 2 | Kaur, Ji et al. 2021 | USA | Acute  +Covid-19 | Recovered  +Covid-19 | 6  4/2 | 6  3/3 | 12  7/5 | 47(14.4) | 36.3(9.4) | Serum | KYN, TRP | UHLC/MS/MS | 1 | 3 | 12 | High TRP , No change in KYN |
| 3 | Lopez-Hernandez, Monarrez-Espino et al. 2021 | Mexico | +Covid-19 | NA | 122  72/50 | 39  18/21 | 161  90/71 | 55.7(10.0) | 43.8(12.3) | Plasma | KYN, TRP | LC–MS/MS | 1 | 5 | 9 | High KYN/TRP ratio |
| 4 | Guo, Schurink et al. 2021 | Netherlands | +Covid-19 | NA | 21  NA | NA | 21  NA | 61.8(29.4) | NA | Plasma | KYN,  TRP, | UPLC-MS/MS | 2 | 3 | 15 | High KYN, Low TRP |
| 5 | Danlos, Grajeda-Iglesias et al. 2021 | France | +Covid-19 | NA | 72  NA | 29  NA | 101  NA | NA | NA | Plasma | TRP, KYN,AA,KA,3HK | UHPLC/MS | 1 | 4 | 8 | Low TRP, High KYN, AA, KA, 3HK across severity |

**ESF. Table 5**. Results of Meta-regression

| Variables | No. of Studies | Covariates | 1-sided p-value | Z-Value |
| --- | --- | --- | --- | --- |
| KYN/TRP | 10 | Medium | 0.047 | 1.67 |
| KA | 4 | Medium | 0.006 | 2.46 |
| KA/KYN+TRP | 7 | Severity | 0.003 | -2.75 |
| KYN+KA/TRP | 10 | Medium | 0.051 | 1.63 |
| KYN/KA | 6 | Total subject | 0.022 | 2.0 |
| KA/KYN | 6 | Total subject | 0.022 | 2.0 |

**ESF, Table 6.** PRISMA checklist

| **Section/topic** | **#** | **Checklist item** | **Reported on page #** |
| --- | --- | --- | --- |
| **TITLE** | | | |
| Title | 1 | Identify the report as a systematic review, meta-analysis, or both. | 1 |
| **ABSTRACT** | | | |
| Structured summary | 2 | Provide a structured summary including, as applicable: background; objectives; data sources; study eligibility criteria, participants, and interventions; study appraisal and synthesis methods; results; limitations; conclusions and implications of key findings; systematic review registration number. | 3 |
| **INTRODUCTION** | | | |
| Rationale | 3 | Describe the rationale for the review in the context of what is already known. | 5 |
| Objectives | 4 | Provide an explicit statement of questions being addressed with reference to participants, interventions, comparisons, outcomes, and study design (PICOS). | 7 |
| **METHODS** | | | |
| Protocol and registration | 5 | Indicate if a review protocol exists, if and where it can be accessed (e.g., Web address), and, if available, provide registration information including registration number. | 8 |
| Eligibility criteria | 6 | Specify study characteristics (e.g., PICOS, length of follow-up) and report characteristics (e.g., years considered, language, publication status) used as criteria for eligibility, giving rationale. | 8 |
| Information sources | 7 | Describe all information sources (e.g., databases with dates of coverage, contact with study authors to identify additional studies) in the search and date last searched. | 8 |
| Search | 8 | Present full electronic search strategy for at least one database, including any limits used, such that it could be repeated. | 9 |
| Study selection | 9 | State the process for selecting studies (i.e., screening, eligibility, included in systematic review, and, if applicable, included in the meta-analysis). | 9 |
| Data collection process | 10 | Describe method of data extraction from reports (e.g., piloted forms, independently, in duplicate) and any processes for obtaining and confirming data from investigators. | 10 |
| Data items | 11 | List and define all variables for which data were sought (e.g., PICOS, funding sources) and any assumptions and simplifications made. | 10 |
| Risk of bias in individual studies | 12 | Describe methods used for assessing risk of bias of individual studies (including specification of whether this was done at the study or outcome level), and how this information is to be used in any data synthesis. | 11 |
| Summary measures | 13 | State the principal summary measures (e.g., risk ratio, difference in means). | 11 |
| Synthesis of results | 14 | Describe the methods of handling data and combining results of studies, if done, including measures of consistency (e.g., I^2^) for each meta-analysis. | 12 |
| Risk of bias across studies | 15 | Specify any assessment of risk of bias that may affect the cumulative evidence (e.g., publication bias, selective reporting within studies). | 12 |
| Additional analyses | 16 | Describe methods of additional analyses (e.g., sensitivity or subgroup analyses, meta-regression), if done, indicating which were pre-specified. | 12 |
| **RESULTS** | | |  |
| Study selection | 17 | Give numbers of studies screened, assessed for eligibility, and included in the review, with reasons for exclusions at each stage, ideally with a flow diagram. | 12 |
| Study characteristics | 18 | For each study, present characteristics for which data were extracted (e.g., study size, PICOS, follow-up period) and provide the citations. | 13 |
| Risk of bias within studies | 19 | Present data on risk of bias of each study and, if available, any outcome level assessment (see item 12). | Table 3, 46 |
| Results of individual studies | 20 | For all outcomes considered (benefits or harms), present, for each study: (a) simple summary data for each intervention group (b) effect estimates and confidence intervals, ideally with a forest plot. | Table 1, 42 |
| Synthesis of results | 21 | Present results of each meta-analysis done, including confidence intervals and measures of consistency. | Table 2, 44 |
| Risk of bias across studies | 22 | Present results of any assessment of risk of bias across studies (see Item 15). | Table 3, 46 |
| Additional analysis | 23 | Give results of additional analyses, if done (e.g., sensitivity or subgroup analyses, meta-regression [see Item 16]). | Table 2, 44 |
| **DISCUSSION** | | |  |
| Summary of evidence | 24 | Summarize the main findings including the strength of evidence for each main outcome; consider their relevance to key groups (e.g., healthcare providers, users, and policy makers). | 18-24 |
| Limitations | 25 | Discuss limitations at study and outcome level (e.g., risk of bias), and at review-level (e.g., incomplete retrieval of identified research, reporting bias). | 24 |
| Conclusions | 26 | Provide a general interpretation of the results in the context of other evidence, and implications for future research. | 24, figure 1 |
| **FUNDING** | | |  |
| Funding | 27 | Describe sources of funding for the systematic review and other support (e.g., supply of data), role of funders for the systematic review. | 25 |

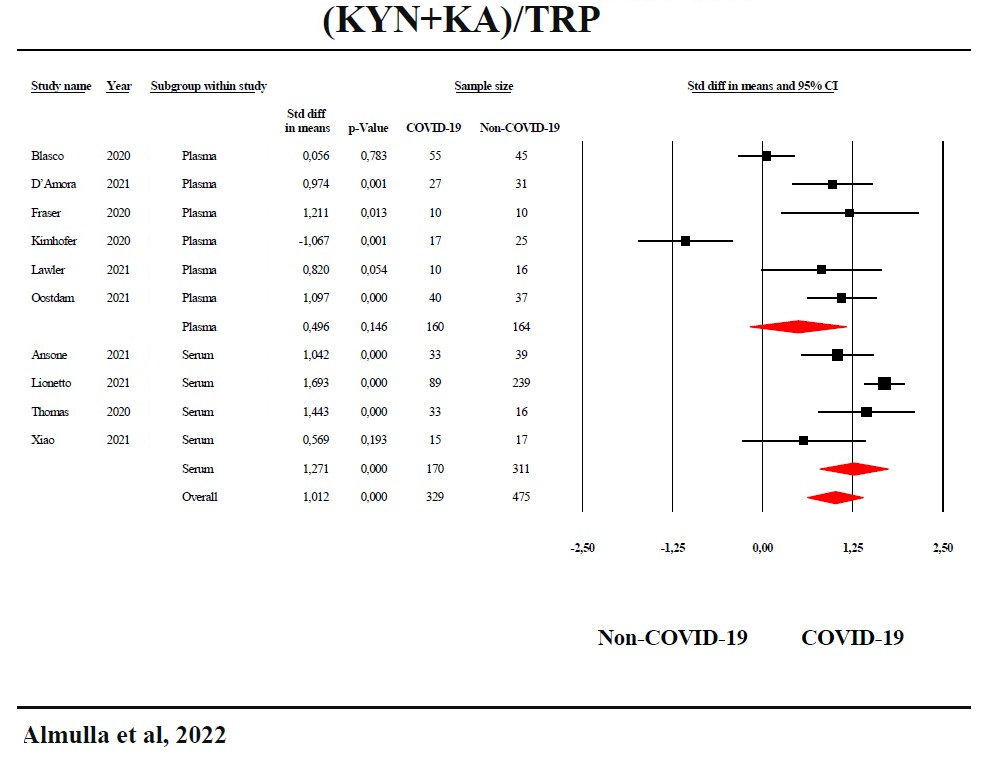

ESF, Figure 1: Forest plot with the results of a meta-analysis performed on the kynurenine + kynurenic acid / tryptophan (KYN+KA)/TRP ratio in COVID-19 patients versus non-COVID-19 controls.

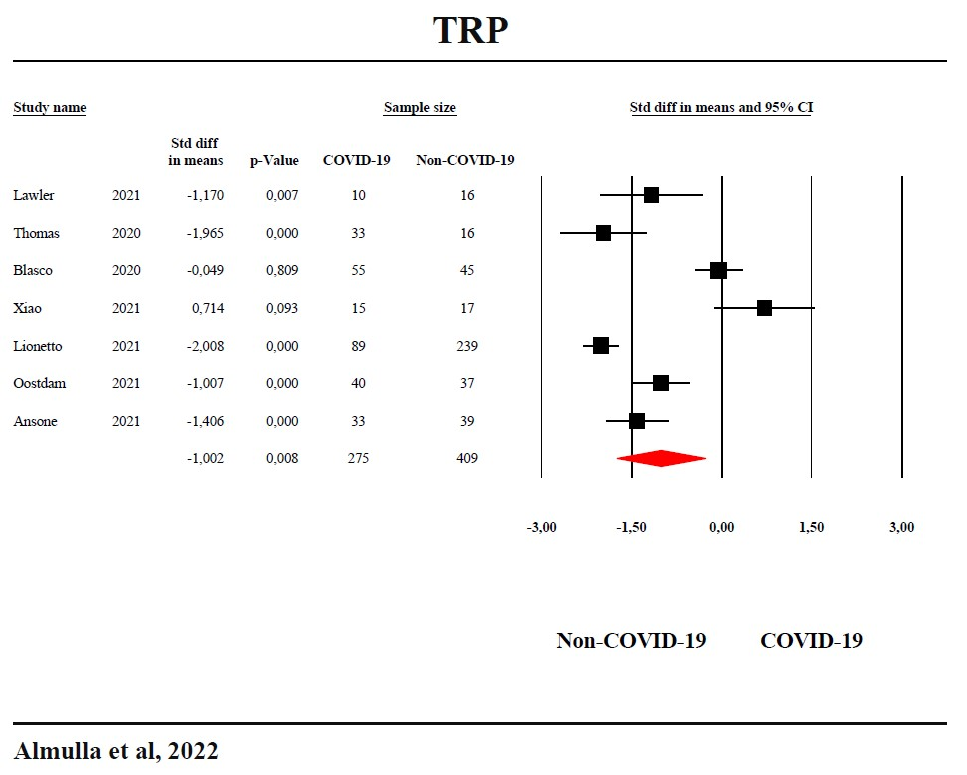

ESF, Figure 2: Forest plot with the results of the meta-analysis performed on tryptophan (TRP) in COVID-19 patients versus non-COVID-19 controls.

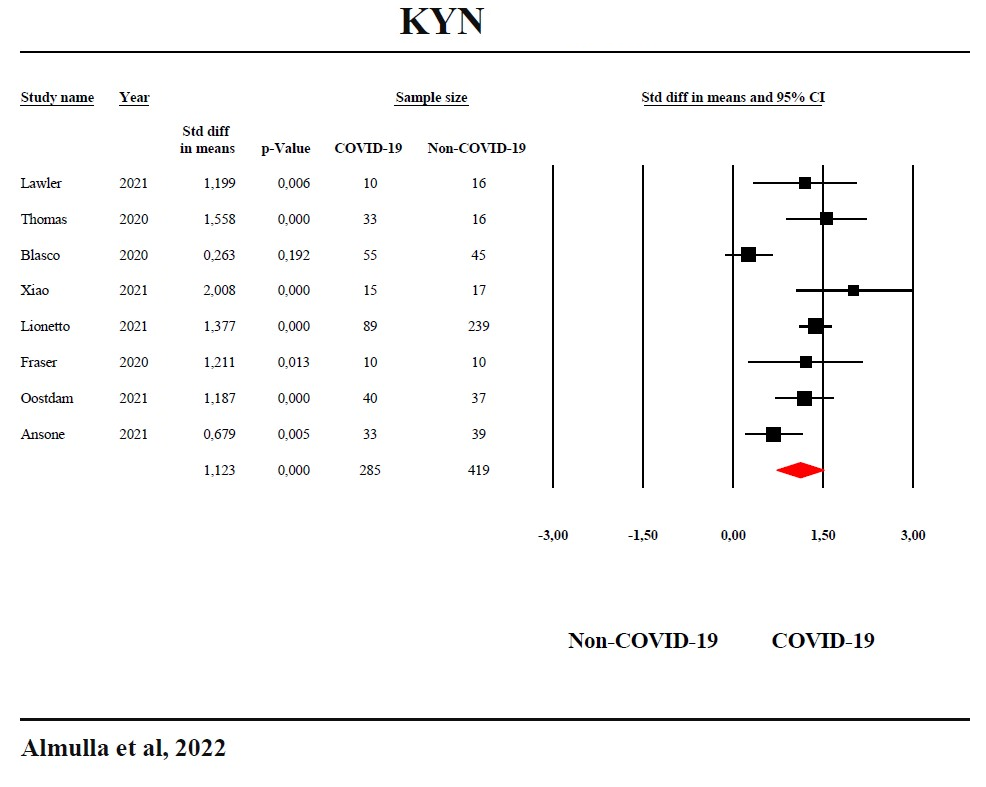

ESF, Figure 3: Forest plot with the results of a meta-analysis performed on kynurenine (KYN) in COVID-19 patients versus non-COVID-19 controls.

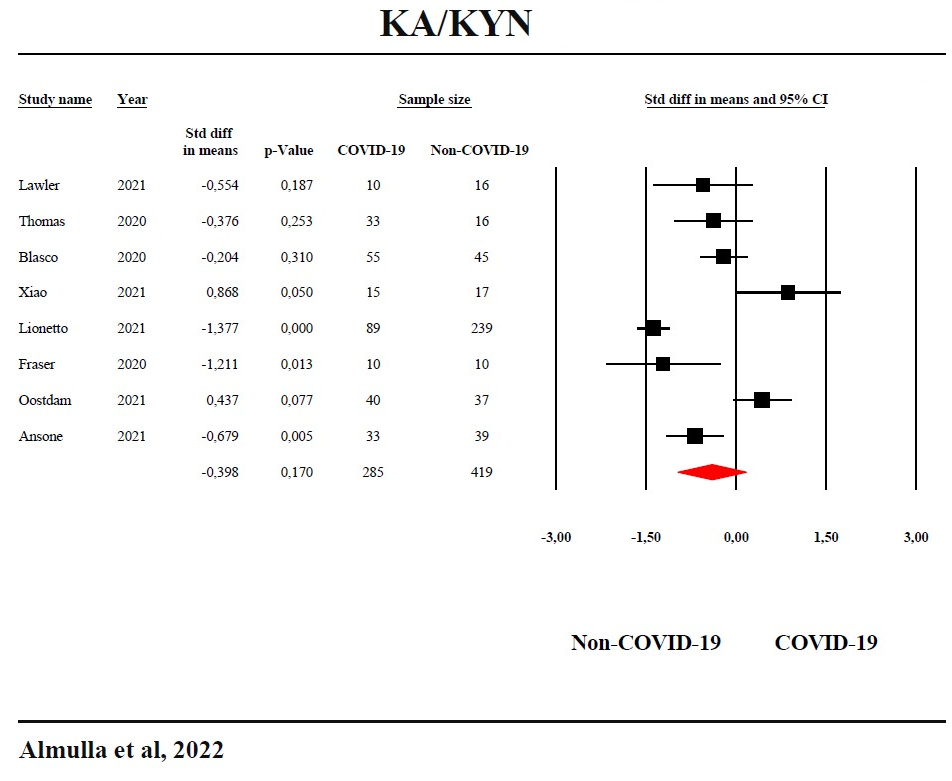

ESF, Figure 4: Forest plot with the results of a meta-analysis performed on the kynurenic acid / kynurenine (KA/KYN) ratio in COVID-19 patients versus non-COVID-19 controls.

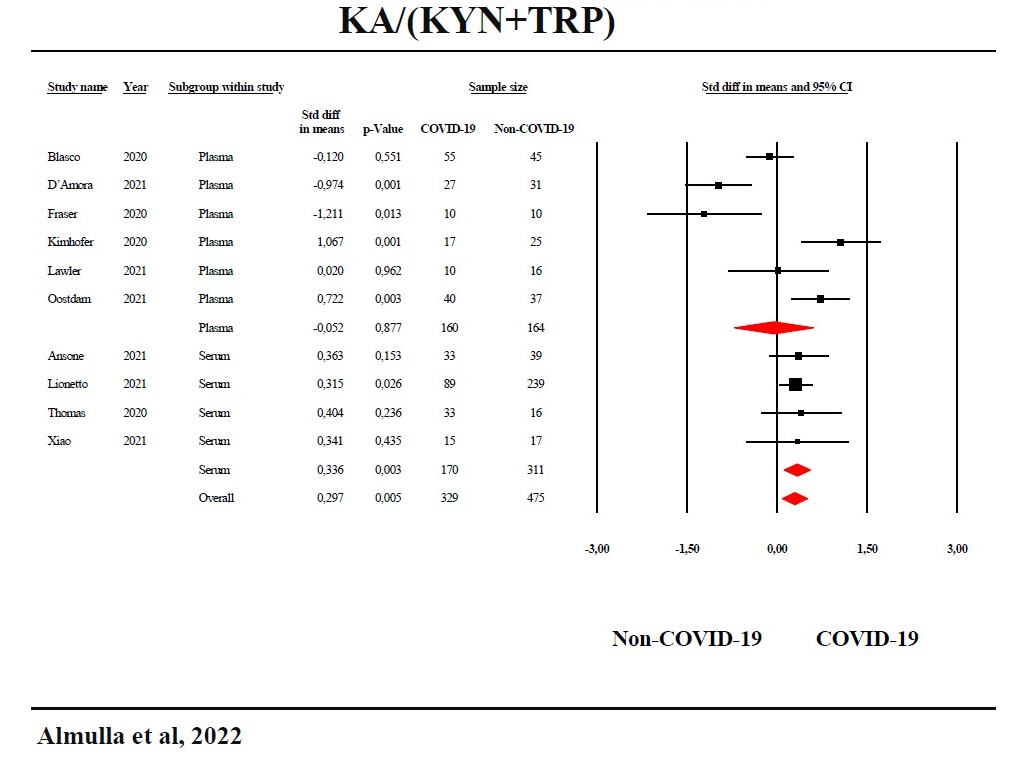

ESF, Figure 5: Forest plot with the results of the meta-analysis performed on the kynurenic acid / kynurenine + tryptophan (KA/KYN+TRP) ratio in COVID-19 patients versus non-COVID-19 controls.

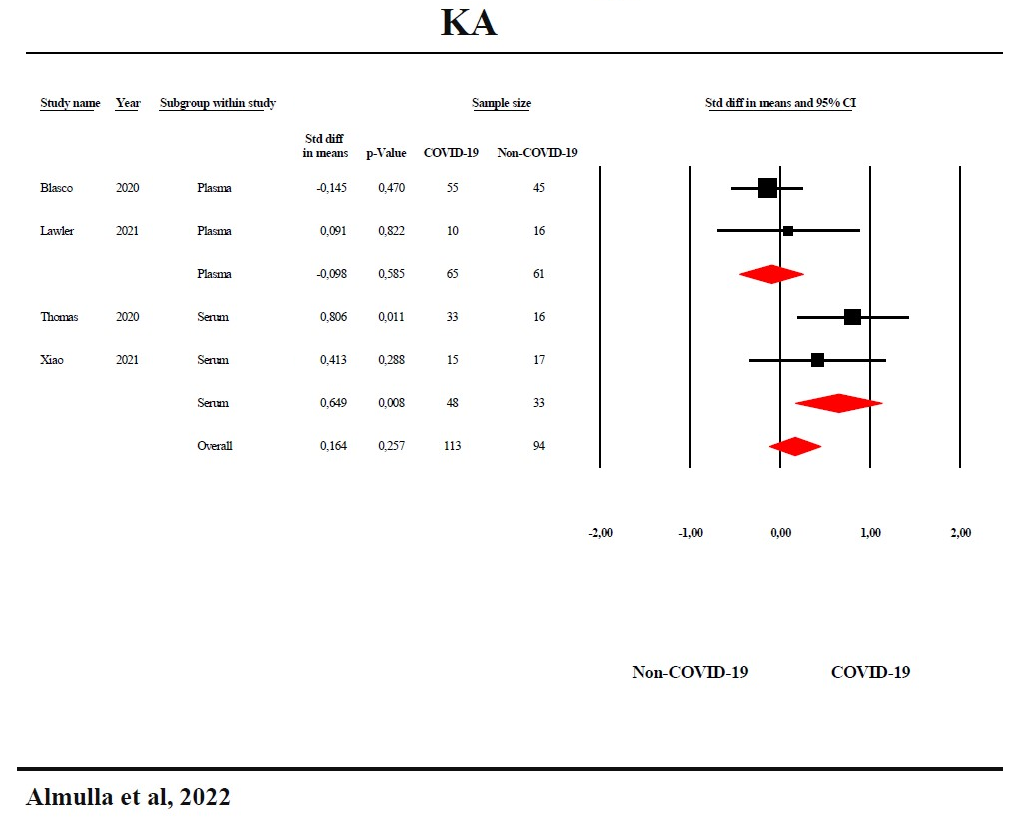

ESF, Figure 6: Forest plot with the results of a meta-analysis performed on kynurenic acid (KA) in COVID-19 patients versus non-COVID-19 controls.

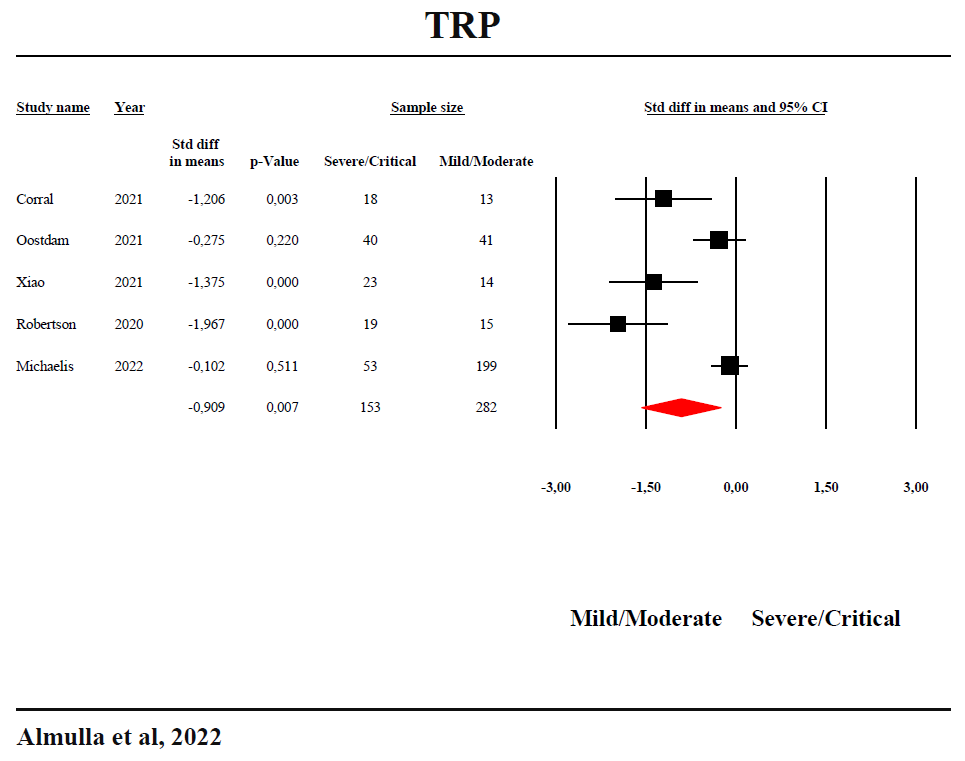

ESF, Figure 7: Forest plot with the results of the meta-analysis performed on tryptophan (TRP) in severe/critical COVID-19 versus mild/moderate COVID-19.

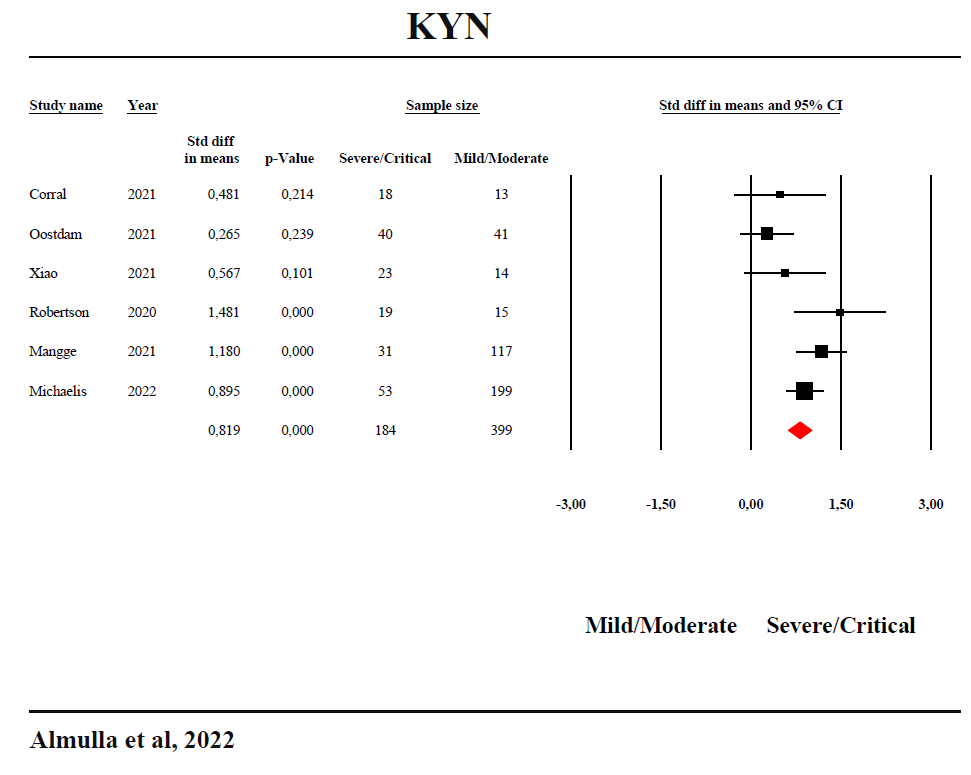

ESF, Figure 8: Forest plot with the results of a meta-analysis performed on kynurenine (KYN) in severe/critical COVID-19 versus mild/moderate COVID-19.

**Abbreviation:**

TRP: Tryptophan

KYN: Kynurenine

KA: Kynurenic acid

3HK: 3-Hydroxykynurenine

AA: Anthranilic acid

3HA: 3-Hydroxyanthranilic acid

XA: Xanthurenic acid

QA: Quinolinic acid

PA: Picolinic acid

IDO: Indoleamine 2,3 dioxygenase

TDO: Tryptophan 2,3 dioxygenase

KAT: Kynurenine aminotransferase

KMO: Kynurenine 3-monooxygenase

KYNU: Kynureninase

NAD^+^: Nicotinamide adenine dinucleotide

COVID-19 : Coronavirus disease-2019

SARS-Cov-2: Severe acute respiratory syndrome coronavirus 2

TRYCATs: Tryptophan Catabolites

TRYCAT pathway: Tryptophan catabolite pathway

KP: Kynurenine pathway

SMD: Standardized mean difference

nCoV: Novel coronavirus

IL-6: Interleukin-6

IFN-γ: Interferon-gamma

CT: Computed tomography

PGI2: Prostaglandin I2

TxA2: Thromboxane A2

O&NS: Oxidative and nitrosative stress

ATP: Adenosine triphosphate

NO: Nitric oxide

NADP: Nicotinamide adenine dinucleotide phosphate

T2DM: Type 2 diabetes mellites

BMI: Body mass index

AhR: Aryl hydrocarbon receptors

NK: Natural Killer cells

CD8+: Cluster of differentiation

MOOSE: Meta-Analyses of Observational Studies in Epidemiology

CSF: Cerebrospinal fluid

RT-PCR: Real time-polymerase chain reaction

SD: Standard definition

IOR: Interquartile range

ICS: Immune confounder scale

CI: Confidence intervals

LC-MS: Liquid chromatography-mass spectrometry

LC-MS/MS: Liquid chromatography with two mass spectrometry

UHPLC-MS: Ultra-high-performance liquid-chromatography- mass spectrometry

LC-HRMS: Liquid chromatography–high-resolution mass spectrometry

LC-UV: Liquid chromatography-UV detection

CKD: Chronic kidney disease

α7nAChr: alpha 7 nicotinic acetylcholine receptor

NMDA: N-methyl-D-aspartate

AMPA: α-amino-3-hydroxy-5-methyl-4isoxazolepropionic acid
